# Phenome-wide and proteomic analysis of APOE alleles across different ancestries in the All of Us Research Program

**DOI:** 10.64898/2026.09.26.26364080

**Authors:** Alejandro Mejia-Garcia, Pablo Valderrama-Carmona, Thomas M. Zheng, Zhenqian Wang, Hsuan Megan Tsao, Jean Jung, Carlos A Orozco, Chen-Yang Su, Sirui Zhou

## Abstract

Apolipoprotein E (APOE) ε4 is the strongest common genetic risk factor for Alzheimer’s disease and a determinant of cardiometabolic risk, yet studies of APOE genotypes with disease associations and plasma proteomic signatures have been primarily restricted to European-ancestry cohorts.

**Methods:** We performed a phenome-wide association study of APOE genotype in 367,757 All of Us participants across six genetic ancestry groups. APOE alleles were tested against 2,632 phenotypes using an ordinal Alzheimer’s-disease-risk hierarchy (ε2/ε2 < ε2/ε3 < ε3/ε3 < ε2/ε4 < ε3/ε4 < ε4/ε4), adjusted for age, sex at birth, EHR length, and 16 genetic principal components. Sex- and ancestry-stratified analyses used Cochran’s Q for heterogeneity. We evaluated ε2 and ε4 dosage effects on plasma protein levels in 9,132 participants across five ancestries.

**Findings:** We identified 27 phenotypes associated with APOE, mainly in neurological and lipid-related traits. Five lipid-related traits showed sex heterogeneity, with larger effects in females. Nine associations showed ancestry-specific effects. APOE ε4 showed stronger protection against fatty liver disease in East Asians with lack of protection in African ancestry; and larger dementia risk for Europeans. Finally, SNAP25 was identified as positively associated with APOE ε4 dosage across ancestries, and CDC42EP1, APOA1 and ITGB5 shown associations with APOE ε2/ε4 specifically in non-European populations.

**Interpretation:** Ancestry-heterogeneous APOE effects, particularly for liver and dementia phenotypes, argue for ancestry-aware interpretation of APOE-based risk. Plasma SNAP25 is a candidate ancestry-portable biomarker of APOE-ε4 effects, and ancestry-specific proteins nominate mechanisms for the observed phenotypic heterogeneity.

**Research in context Evidence before this study:** The APOE ε4 variant is an established genetic risk factor for hyperlipidemia, Alzheimer’s disease, dementia, ischemic heart disease and, more recently, non-alcoholic fatty liver disease. Prior phenome-wide association studies (PheWAS) of APOE have been conducted largely in European-ancestry cohorts. A recent APOE PheWAS in the All of Us Research Program (n = 181,880) identified 17 Bonferroni-significant phenotypes and preliminary evidence of ancestry heterogeneity for hyperlipidemia and mild cognitive impairment, but statistical power in non-European groups limited systematic characterization of ancestry-specific effects. Recent plasma proteomic studies have described APOE-associated protein signatures shared with neurodegenerative diseases, but these were confined to European-ancestry participants, leaving open whether such signatures generalize across ancestries.

**Added value of this study:** Using the All of Us v9 release, we approximately doubled the sample size of the prior APOE PheWAS in this cohort (n = 367,757 across six ancestry groups, including ∼146,000 non-European participants), enabling the first well-powered assessment of ancestry heterogeneity of APOE effects. We identify 27 Bonferroni-significant phecodes, 11 novel compared to the previous APOE PheWAS in All of Us — including vascular dementia, dementia with Lewy bodies, and angina pectoris. We detect significant sex heterogeneity for five lipid-related phecodes and significant ancestry heterogeneity for nine phecodes, including stronger protection against fatty liver and chronic liver disease in East Asian participants and larger dementia risk in European participants. Complementing the phenotypic analysis, we present the first ancestry-stratified plasma proteomic characterization of APOE genotypes (n = 9,132), identifying SNAP25 as a robust APOE-ε4–associated protein across ancestries and ancestry-specific signals for IL32, CDC42EP1, APOA1, and ITGB5.

**Implications of all the available evidence:** APOE-associated risk estimates derived predominantly from European-ancestry data may not transfer directly to other populations, and our findings support incorporating ancestry into the interpretation and calibration of APOE-based risk prediction. The ancestry-heterogeneous protection against liver disease and the parallel APOA1 signal absent in East and South Asian participants together offer a mechanistic hypothesis linking APOE-driven lipid partitioning to hepatic outcomes. Plasma SNAP25 is proposed as an accessible, ancestry-portable biomarker of APOE-ε4 effects and Alzheimer’s disease progression. Ancestry-specific proteins (IL32 and CDC42EP1 in African ancestry, ITGB5 in Admixed American ancestry) constitute potential drivers of ancestry heterogeneity in APOE-related disease risk.

## Introduction

Apolipoprotein E (APOE) is a lipid-binding protein with a central role in lipid metabolism through its interaction with the low-density lipoprotein receptor (LDLR) and related receptors ^1^. Three common APOE alleles have been described — ε2, ε3, and ε4 — defined by two single-nucleotide polymorphisms (rs429358 and rs7412) that alter amino acids at positions 112 and 158 of the mature protein ^2^. These substitutions alter the interaction of each isoform with the LDL receptor. APOE ε2 binds LDLR with less affinity, impairing clearance of APOE-containing remnant lipoproteins, resulting in the compensatory upregulation of hepatic LDLR, which lowers plasma LDL-C ^3^. APOE ε4 promotes faster catabolism of triglyceride-rich remnants, driving downregulation of LDLR and higher plasma LDL-C ^4^.

The ε4 allele is the strongest common genetic risk factor for late-onset Alzheimer’s disease (AD) ^5,6^, with each additional copy associated with earlier age of onset and increased dementia risk, whereas ε2 is considered protective ^7–9^. Beyond neurological outcomes, APOE variation has been linked to atherosclerosis, stroke, hyperlipidemia, and non-alcoholic fatty liver disease, among other conditions ^4,10–14^. Ancestry-specific allele frequency patterns are well documented: ε4 frequency is highest in African-ancestry populations ^11^, consistent with evolutionary evidence that ε4 is the ancestral allele and that ε3 and ε2 emerged and rose in frequency under positive selection during human population expansion ^15,16^. However, whether APOE confers the same phenotypic and molecular risk across ancestries — a prerequisite for equitable clinical use — remains largely unknown, because the vast majority of APOE studies have been conducted in European-ancestry cohorts.

Phenome-wide association studies (PheWAS) provide a systematic framework for characterizing the pleiotropic effects of APOE alleles across electronic health record (EHR)-derived phenotypes and enable stratified analyses by sex and ancestry. Khajouei et al. (2025) recently reported a PheWAS of APOE in 181,880 All of Us participants from six different ancestries, identifying 17 Bonferroni-significant associations and providing evidence for ancestry heterogeneity in hyperlipidemia and mild cognitive impairment ^11^. However, their findings in non-European ancestry groups were limited by sample size. Whether APOE risk transfers across populations for the disease outcomes most relevant to clinical use remains unresolved.

Recent large-scale proteomic studies also characterized the blood proteomic signatures of APOE variation. Lu et al. (2026) performed a multi-cohort, multi-platform analysis in five cohorts mainly from European participants (GNPC, BioFINDER-2, ADNI, UK Biobank, PPMI), integrating SomaLogic and Olink Explore plasma and CSF data to identify ε4- and ε2-associated protein signatures spanning lipid metabolism, immune regulation, and pathways shared across neurodegenerative diseases ^17^. A parallel study reported that APOE ε4 carriers share immune-related proteomic changes across neurodegenerative conditions ^18^. APOE-associated protein alterations can be detected before amyloid-β pathology and remain stable across age and disease progression, supporting their utility as early biomarkers ^17^. However, both studies consisted of mainly European-ancestry participants, and the largest Olink plasma dataset in Lu et al. (UK Biobank, N=4,813, Olink Explore 3,072) is a subset of the newer Olink Explore HT panel available in All of Us. Whether these signatures generalize across ancestries and whether ancestry-specific proteomic effects exist remain unresolved.

This study aims to address abovementioned gaps by testing whether APOE-related risk generalizes across ancestries at both the phenotypic and blood proteomic level, leveraging the recent released data from All of Us. We will (i) replicate and extend prior APOE PheWAS findings across 367,757 participants and 2,632 phecodes from All of Us Research Program v9 release, (ii) test effect heterogeneity across sex and ancestry with increased power, and (iii) characterize the ancestry-stratified plasma proteomic signature of APOE variation as a step toward mechanistic understanding. We hypothesize that the expanded sample size and multi-ancestry design will reveal ancestry-heterogeneous APOE effects at both the phenotypic and plasma proteomic level that are undetectable in prior European-ancestry–dominated studies.

## Methods

### The All of Us research program

The All of Us Research Program (AoU) is a diverse cohort of individuals from different ancestry groups, including Europeans, Africans, admixed Americans (Latin Americans) and Asians (South and East Asians) ^19^. All participants consented to be part of the study and provided access to their electronic health record (EHR).

## Ethics

Access to All of Us received institutional approval by McGill University (IRB: 2107073). This study did not directly involve the recruitment of human participants. We accessed data from All of Us participants using the cloud Workbench 2.0. We followed all the guidelines and restrictions given by the All of Us team.

### Study cohort

We analyzed participants of the All of Us Research Program (v9 Curated Data Repository release) with short-read whole-genome sequencing (srWGS) data, EHR and inferred genetic ancestry. Sex at birth was restricted to Male or Female. This yielded a primary analytic cohort of 367,757 participants across six ancestry groups: European (EUR), African (AFR), Admixed American (AMR), East Asian (EAS), South Asian (SAS), and Middle Eastern (MID) as defined by All of Us gnomAD-based ancestry inference.

### Genetic ancestry estimation

Genetic ancestry groups were predicted by the Data and Research Center (DRC) with a random forest classifier trained on 16 genetic principal components (PCs), using the 1000 genomes project and Human Genome Diversity Project (HGDP) as reference ^19^. In detail, shared sites between the All of Us cohort and the reference panel (1KG and HGDP) were used. Only autosomal and biallelic SNPs with a frequency > 0.1% and a call rate > 99% were kept. An R^2^ = 0.1 was used to determine variants in linkage disequilibrium, keeping 130,660 independent SNPs.

Genetic PCs were then derived using the hwe_normalized_pca function in the Hail Python package. After, a random forest classifier was trained on the reference panel using this set of high-quality variants and 16 genetic PCs ^19^.

### APOE genotyping

APOE genotypes were determined from Whole-genome sequencing (WGS) at rs429358 and rs7412. SNP data was retrieved from the All of Us WGS, specifically from the Exome subset (extracted from the whole genome) which contains only coding variants for 535,000 participants. Genotype extraction was performed using PLINK 2.0. APOE genotypes were assigned based on allele combinations on these two SNPs and an in-house Python script (Supplementary Table 1). Allele and genotype frequencies were estimated for the whole cohort and by ancestry and sex.

### Phenotype ascertainment

Phenotypes (phecodes) were generated from ICD-9-CM and ICD-10-CM codes in the AoU OMOP database using the PheWAS ToolKit (PheTK) v.0.3.3, applying phecodeX (3,517 phecodes) and requiring ≥2 code occurrences for a case-positive definition. This yielded 32,108,019 person-phecode events across 367,,757 participants.

### PheWAS

A logistic regression was performed for each phecode with the ordinal APOE variable as the exposure of interest, using the following order given by Khajouei et al. 2025 based on Alzheimer’s disease risk hierarchy: ε2/ε2 (0) < ε2/ε3 (1) < ε3/ε3 (2) < ε2/ε4 (3) < ε3/ε4 (4) < ε4/ε4 (5). The PheWAS ToolKit (PheTK) package was utilized in the All of Us Researcher Workbench (https://workbench.researchallofus.org). Covariates were added using the add.covariates function in the PheTK package. Analysis was adjusted for age, sex at birth, EHR length (duration between first and last EHR event), and the first 16 genetic principal components (PCs). A final cohort of 367,757 was retained after removing missing values for covariates. Phecodes with fewer than 50 cases were excluded, yielding 2,632 tested phecodes and a Bonferroni-corrected significance threshold of α = 0.05/2,632 = 1.90×10⁻ .

### Sex-stratified PheWAS

To evaluate if the associations were consistent across sexes, we repeated the primary analysis but stratified by sex: male (n=142,841) and female (n=224,916) participants. APOE effect heterogeneity was tested per significant phecode from the primary analysis using a Cochran’s Q test as implemented in the statsmodels python package. To confirm sex heterogeneity, we applied a logistic regression adding an interaction term: APOE*Sex.

### Ancestry-stratified PheWAS

To evaluate the strength of the associations per ancestry group, we performed a PheWAS stratified by ancestry using the ancestry labels provided by All of Us. After removing individuals with missing covariates, the following sample sizes were retained for each ancestry group: EUR n=221,901, AMR n=66,856, AFR n=64,261, EAS n=9,549. Cross-ancestry effect heterogeneity was tested per significant phecode using Cochran’s Q statistic, computed using the statsmodels Python package. We performed the analysis using the four largest (EUR/AFR/AMR/EAS), and the three largest (EUR/AFR/AMR) as sensitivity, to rule out that low sample size from EAS was driving the heterogeneity. Phecodes with a false discovery rate (FDR)-significant heterogeneity in both analyses were considered robust.

### Plasma proteomics

Plasma proteomic profiles from AoU v9 Olink HT (batch-normalized, replicates removed; n=9,841 individuals and 5,416 proteins) were used to test APOE allele dosage effects on plasma protein levels. Regression analyses of plasma protein inverse-rank normalized NPX values and ε4 and ε2 allele dosage were performed in a joint model adjusted for age, sex at birth, plate ID, mean NPX value per participant and the first 16 genetic PCs. A final cohort of 9,132 individuals with all covariates was included, and 5,415 proteins were retained (excluding APOE). We analyzed the APOE protein as a positive control, as variants to define APOE genotypes have been shown to alter protein levels ^17^. We then performed ancestry stratified analysis using the same joint model and covariates for EUR (n=2,648), AFR (n=1,439), AMR (n=2,033), EAS (n=1,374) and SAS (n=1,187). We excluded MID ancestry (n= 451) due to low sample size. For each protein that was only significant in one ancestry, we performed a heterogeneity test between the five ancestries as a sensitivity analysis. Selected proteins were also tested for their association with risk for relevant diseases.

### Statistics

Using the PheTK Python package v.0.3.3 in the All of Us Workbench, we applied a logistic regression using APOE genotype as an ordinal variable as previously described. For this model, each OR should be interpreted as an increase in this ordinal order. APOE allelic variation was the independent variable for each available Phecode adjusted by age, sex, and EHR length. In addition, we included 16 genetic PCs generated by All of Us to account for population structure. Stratified analysis by sex and ancestry included the same design and covariate adjustment. In addition, we performed analysis in the overall cohort but with increasing copies of ε2 (ε3/ε3, ε2/ε3, ε2/ε2) and ε4 (ε3/ε3, ε3/ε4, ε4/ε4). To compare with previous literature, we performed an additional analysis by diplotypes (ε2/ε3, ε2/ε2, ε3/ε4, ε4/ε4), comparing against ε3/ε3 (reference). As a sensitivity analysis, we performed the overall PheWAS but including social determinants of health (SDOH) variables such as education levels and social deprivation index (SDI). Heterogeneity testing between sex and ancestry was performed using a Cochran’s Q test, as implemented in the statsmodels package v. 0.14.6. P values were corrected using the FDR method and the significant threshold for heterogeneity was defined as <0.05. Sensitivity analyses were performed using logistic regressions, as implemented in the statsmodels Python package, including associations per diplotype, effect of covariates in significant Phecodes. Interaction analyses were performed by adding a product term APOE*covariate, and the interaction P value was corrected using FDR when multiple comparisons were performed. For the proteomics analysis, we fit a linear regression using the NPX values of each protein as outcome, adjusted by 16 genetic PCs, age, sex, plate ID and the mean NPX value per participant, as recommended elsewhere ^17^. SNAP25 levels between different neurological phenotypes were adjusted by the same covariates as the APOE dosage models. Heterogeneity between ancestries was tested as explained before. We used the STREGA reporting guidelines in the development of this manuscript ^20^.

## Results

### Cohort characteristics

The cohort with genetic, phenotypic and EHR data was comprised of 367,757 participants (Table 1), with 60.3% EUR (N= 221,901), 18.2% AMR (N=66,856), 17.5% AFR (N=64,261), 2.6% EAS (N=9,549), 1.0% SAS (N=3,677), and 0.4% MID (1,513).

**Table 1.**
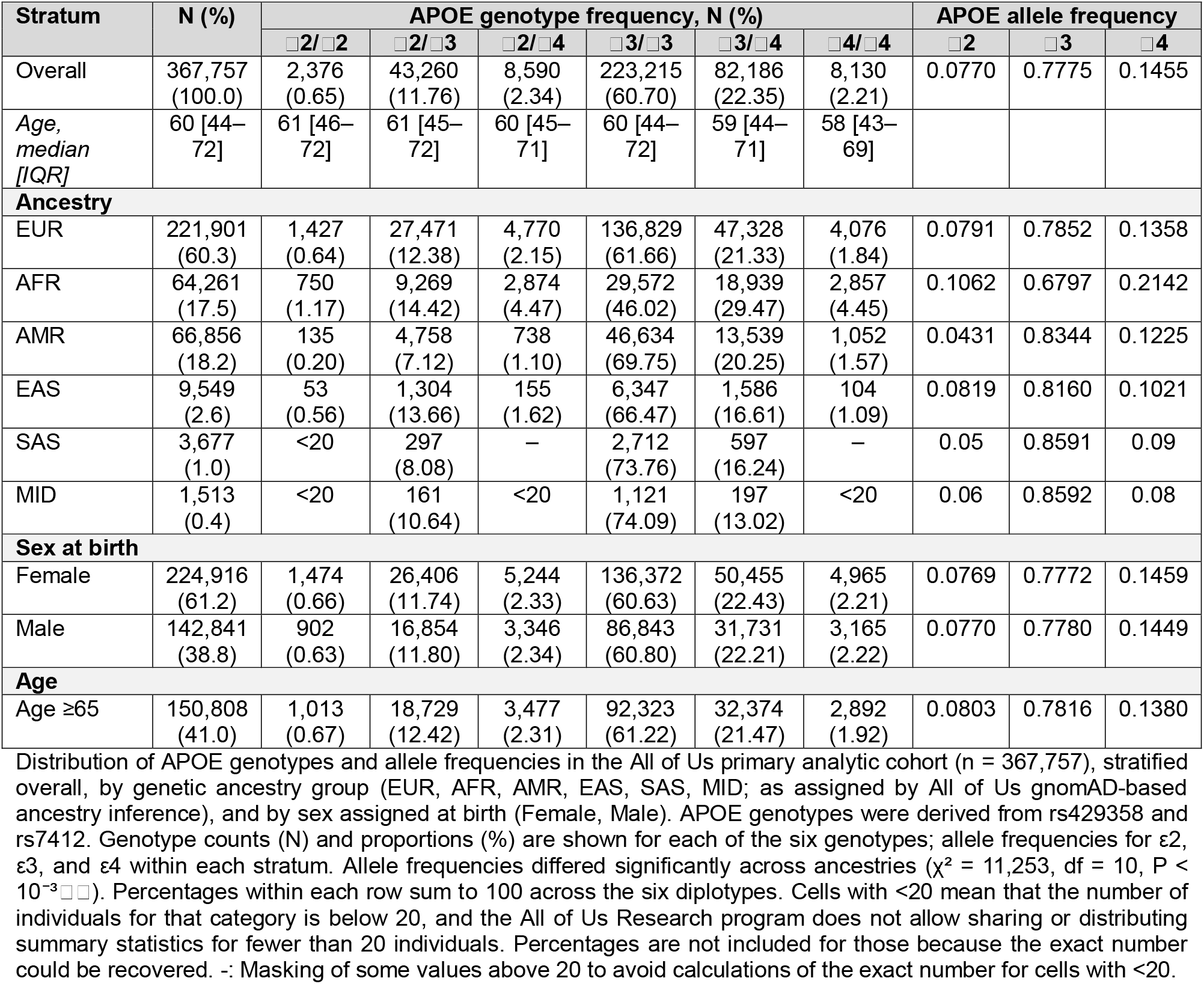
Cohort characteristics by APOE genotype and Allele frequency

| Stratum | N (%) | APOE genotype frequency, N (%) |  |  |  |  |  | APOE allele frequency |  |  |
|---|---|---|---|---|---|---|---|---|---|---|
|  |  | □2/□2 | □2/□3 | □2/□4 | □3/□3 | □3/□4 | □4/□4 | □2 | □3 | □4 |
| Overall | 367,757 (100.0) | 2,376 (0.65) | 43,260 (11.76) | 8,590 (2.34) | 223,215 (60.70) | 82,186 (22.35) | 8,130 (2.21) | 0.0770 | 0.7775 | 0.1455 |
| *Age, median [IQR]* | 60 [44–72] | 61 [46–72] | 61 [45–72] | 60 [45–71] | 60 [44–72] | 59 [44–71] | 58 [43–69] |  |  |  |
| **Ancestry** |  |  |  |  |  |  |  |  |  |  |
| EUR | 221,901 (60.3) | 1,427 (0.64) | 27,471 (12.38) | 4,770 (2.15) | 136,829 (61.66) | 47,328 (21.33) | 4,076 (1.84) | 0.0791 | 0.7852 | 0.1358 |
| AFR | 64,261 (17.5) | 750 (1.17) | 9,269 (14.42) | 2,874 (4.47) | 29,572 (46.02) | 18,939 (29.47) | 2,857 (4.45) | 0.1062 | 0.6797 | 0.2142 |
| AMR | 66,856 (18.2) | 135 (0.20) | 4,758 (7.12) | 738 (1.10) | 46,634 (69.75) | 13,539 (20.25) | 1,052 (1.57) | 0.0431 | 0.8344 | 0.1225 |
| EAS | 9,549 (2.6) | 53 (0.56) | 1,304 (13.66) | 155 (1.62) | 6,347 (66.47) | 1,586 (16.61) | 104 (1.09) | 0.0819 | 0.8160 | 0.1021 |
| SAS | 3,677 (1.0) | <20 | 297 (8.08) | – | 2,712 (73.76) | 597 (16.24) | – | 0.05 | 0.8591 | 0.09 |
| MID | 1,513 (0.4) | <20 | 161 (10.64) | <20 | 1,121 (74.09) | 197 (13.02) | <20 | 0.06 | 0.8592 | 0.08 |
| **Sex at birth** |  |  |  |  |  |  |  |  |  |  |
| Female | 224,916 (61.2) | 1,474 (0.66) | 26,406 (11.74) | 5,244 (2.33) | 136,372 (60.63) | 50,455 (22.43) | 4,965 (2.21) | 0.0769 | 0.7772 | 0.1459 |
| Male | 142,841 (38.8) | 902 (0.63) | 16,854 (11.80) | 3,346 (2.34) | 86,843 (60.80) | 31,731 (22.21) | 3,165 (2.22) | 0.0770 | 0.7780 | 0.1449 |
| **Age** |  |  |  |  |  |  |  |  |  |  |
| Age ≥65 | 150,808 (41.0) | 1,013 (0.67) | 18,729 (12.42) | 3,477 (2.31) | 92,323 (61.22) | 32,374 (21.47) | 2,892 (1.92) | 0.0803 | 0.7816 | 0.1380 |

Distribution of APOE genotypes and allele frequencies in the All of Us primary analytic cohort (n = 367,757), stratified overall, by genetic ancestry group (EUR, AFR, AMR, EAS, SAS, MID; as assigned by All of Us gnomAD-based ancestry inference), and by sex assigned at birth (Female, Male). APOE genotypes were derived from rs429358 and rs7412. Genotype counts (N) and proportions (%) are shown for each of the six genotypes; allele frequencies for ε2, ε3, and ε4 within each stratum. Allele frequencies differed significantly across ancestries (χ<sup>2</sup> = 11,253, df = 10, P < 10<sup>−3□□</sup>). Percentages within each row sum to 100 across the six diplotypes. Cells with <20 mean that the number of individuals for that category is below 20, and the All of Us Research program does not allow sharing or distributing summary statistics for fewer than 20 individuals. Percentages are not included for those because the exact number could be recovered. -: Masking of some values above 20 to avoid calculations of the exact number for cells with <20.

The average age of participants was 58.2 (SD = 16.9) years. Females were younger, with an average age of 56.5 (SD = 16.8) compared to 60.9 (SD = 16.8) in males. In the whole cohort, the most frequent APOE allele was ε3, with an allele frequency of 0.777, compared to 0.077 for the ε2 allele and 0.145 for the ε4 allele. We found significant differences in allele frequencies between ancestries (χ^2^ = 11,253, df = 10, p < 10^-300^), with AFR ancestry displaying the highest frequency of ε4 (0.21) and the lowest ε3 frequency (0.679).

## Primary PheWAS

Of the 3,517 phecodes extracted, 2,632 had at least 50 cases and were included in the analysis. PheWAS of the entire cohort found 27 phecodes associated with ordinal APOE variable (p < 1.90×10⁻ ), spanning cardiovascular, neurological, endocrine/metabolic, gastrointestinal, genetic and mental categories. The most significant associations were with hyperlipidemia (OR 1.15, P = 6.6×10⁻² ) and hypercholesterolemia (OR 1.12, P = 5.4×10⁻¹ ²). The largest effect sizes were observed for Alzheimer’s disease (OR 1.67, P = 9.3×10⁻ ) and dementias (OR 1.31, P = 9.9×10⁻ ). Additional significant associations were observed, such as increased risks for mixed hyperlipidemia, memory loss, mild cognitive impairment, coronary atherosclerosis, ischemic heart disease, vascular dementia as well as decreased risk for chronic liver disease and fatty liver disease (Figure 1, Table 2). These associations are consistent with APOE’s known pleiotropy and involvement in lipid and neurological traits ^11^.

**Figure 1.**
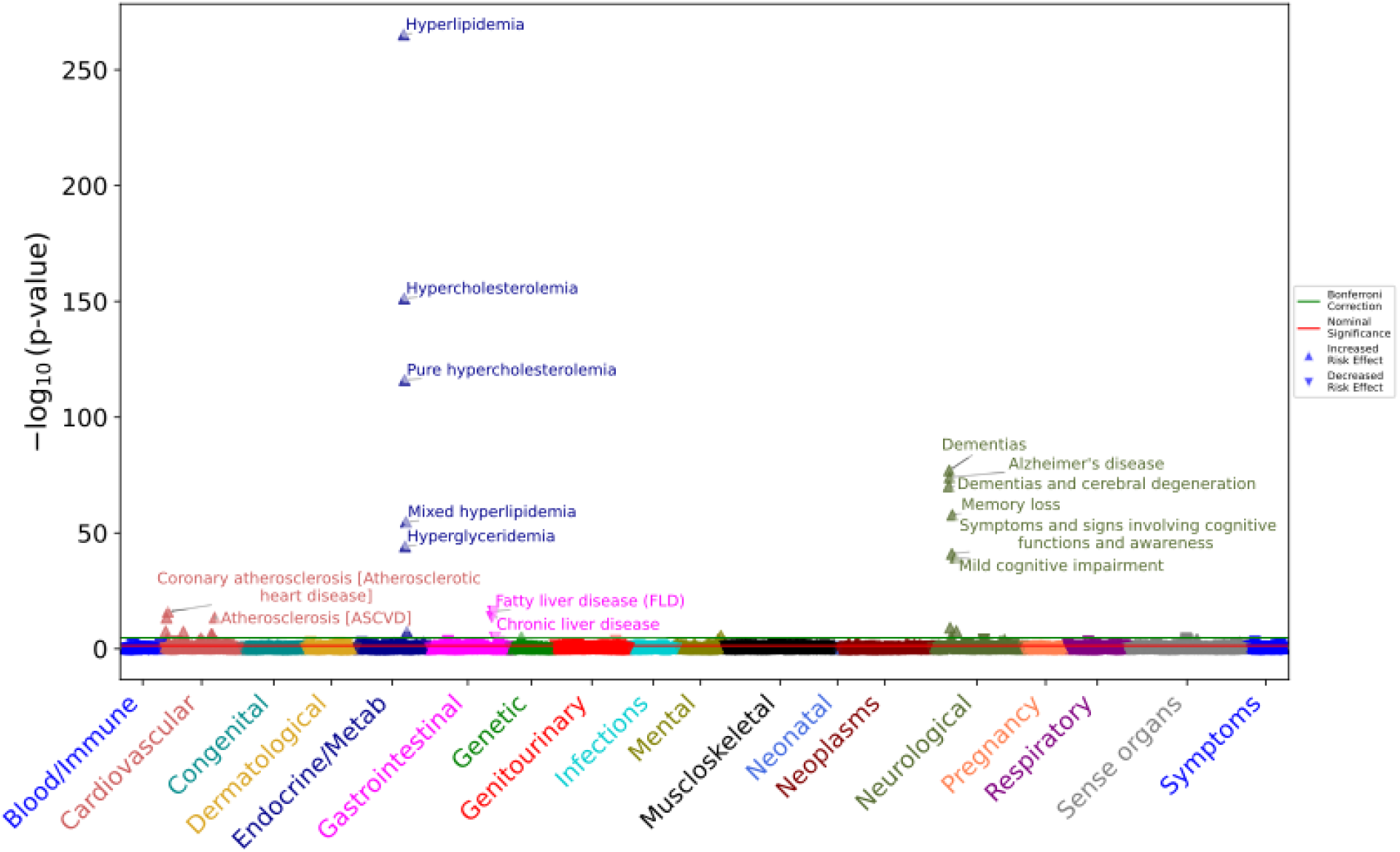
PheWAS analysis of APOE genotypes. APOE genotypes PheWAS analysis in the whole cohort (n= 367,757). Genotypes were arranged hierarchically from ε2 to ε4 (ε2/ε2<ε2/ε3<ε3/ε3<ε2/ε4<ε3/ε4<ε4/ε4). Logistic regressions for each phecode were adjusted by age, sex, 16 genetic PCs and EHR length. A total of 2,632 phecodes were evaluated. The green line represents the significance threshold after Bonferroni correction (p < 1.90×10⁻ ). The red line represents the nominal significance threshold of 0.05. The direction of the triangles represents the direction of the association, with a triangle pointing up meaning an OR > 1 and an upside-down triangle meaning an OR < 1. X-axis shows phecodes aggregated into 18 categories, as implemented by the PheTK package. The y-axis displays the -log10(p-value) for each phecode.

**Table 2.**
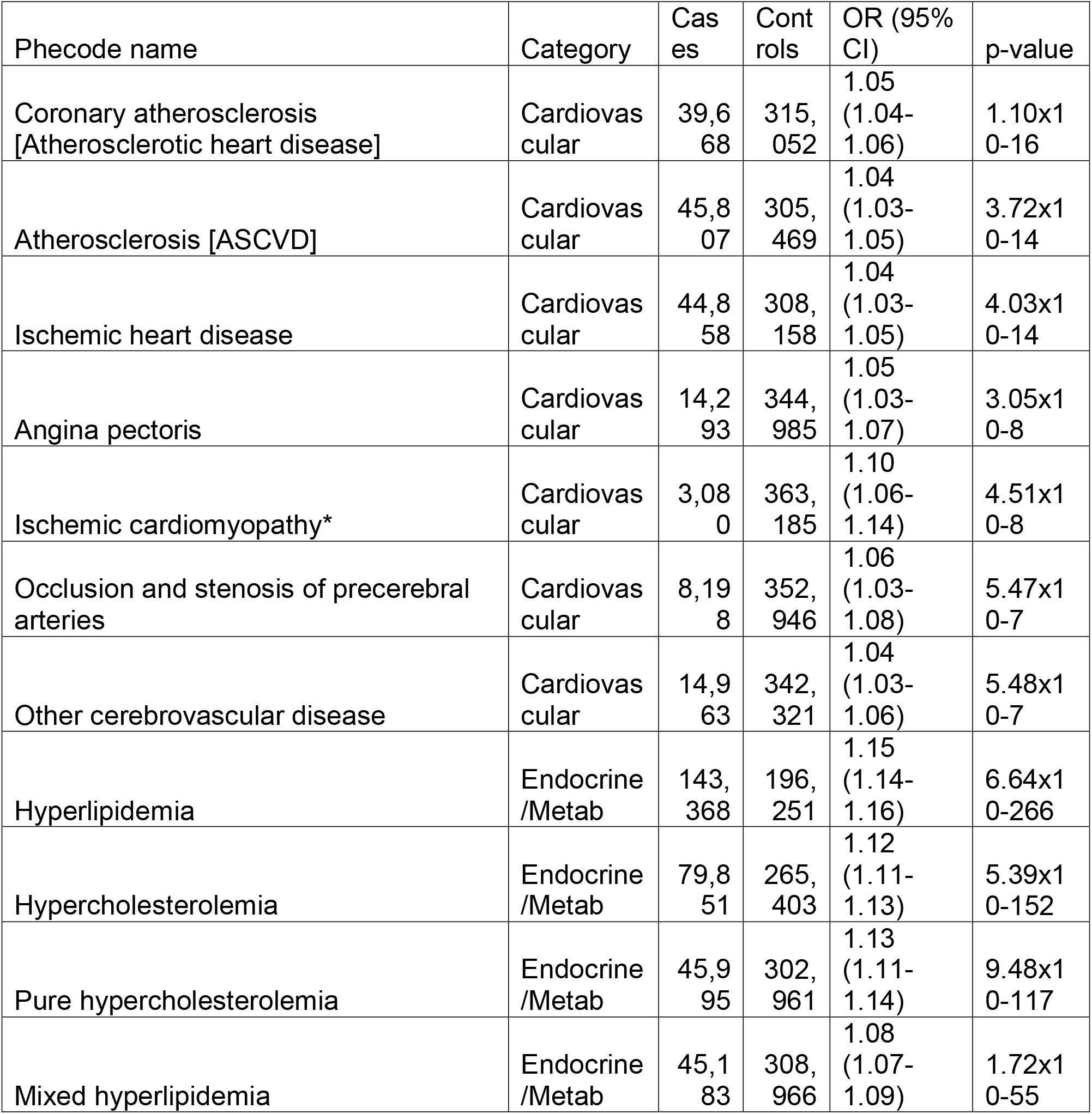
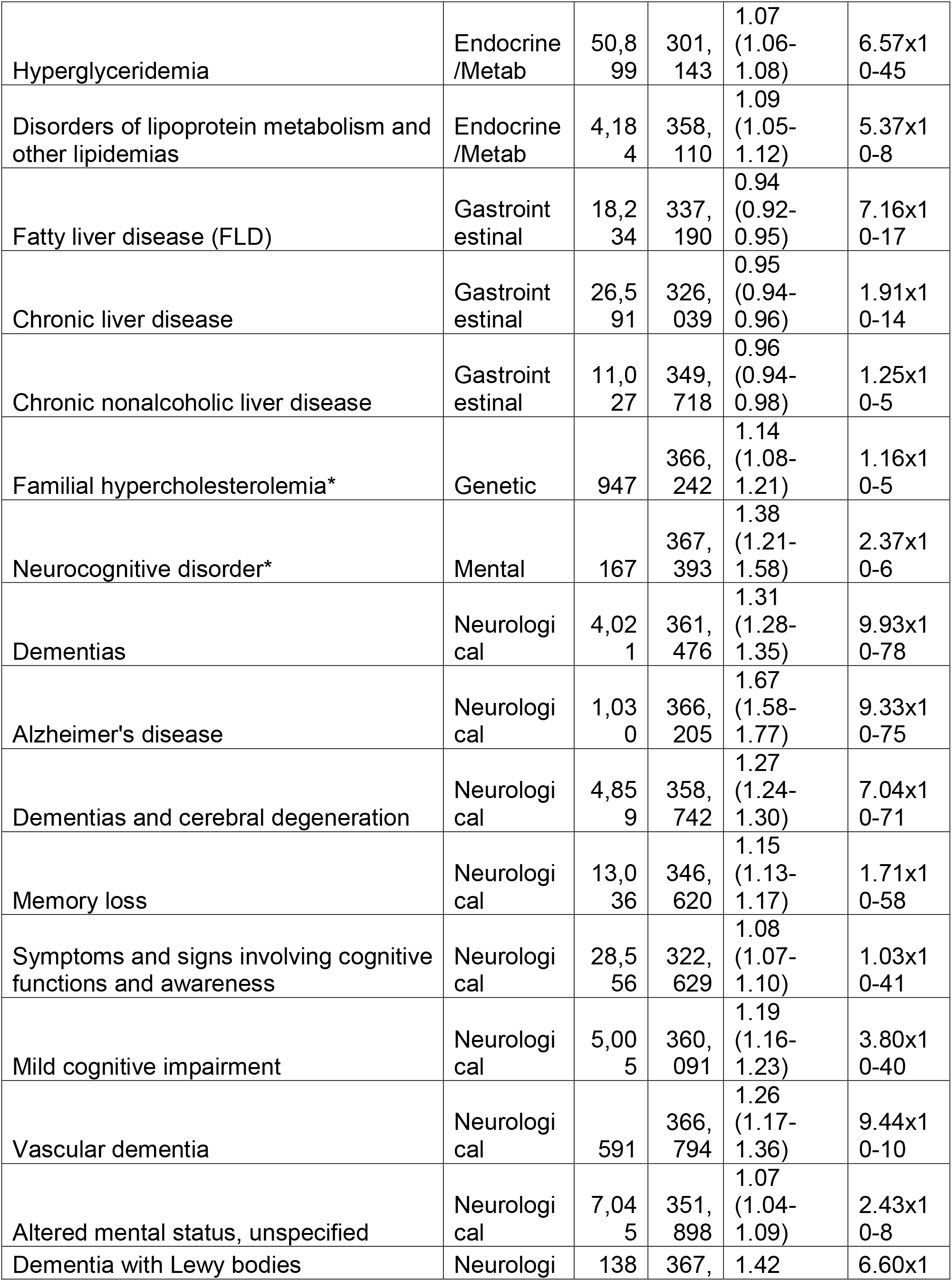
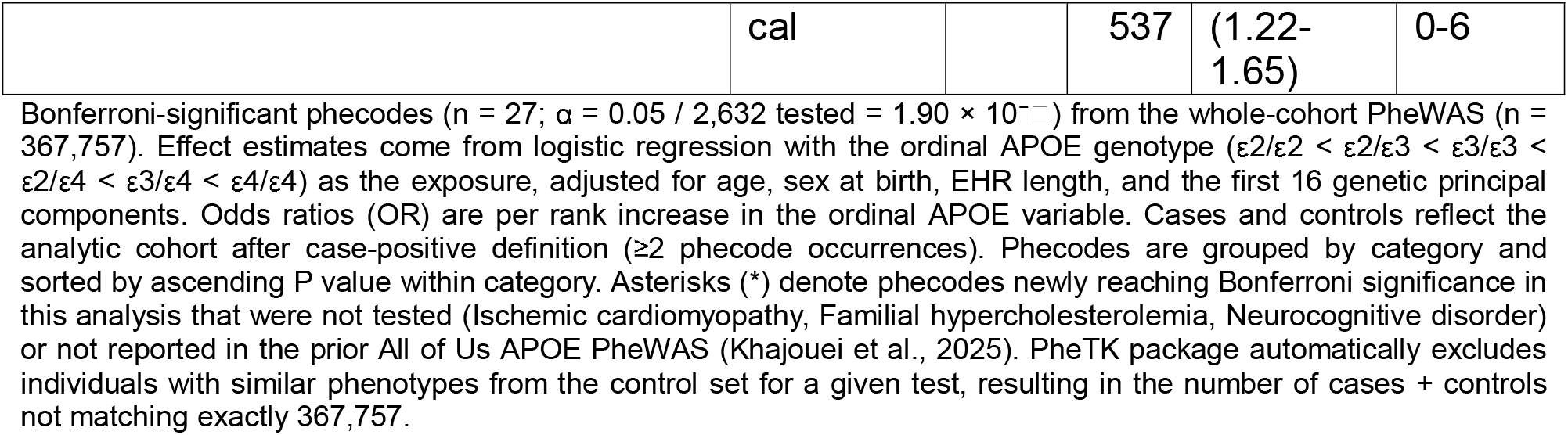
Significant associations between APOE genotypes and 27 phecodes.

| Phecode name | Category | Cases | Controls | OR (95% CI) | p-value |
|---|---|---|---|---|---|
| Coronary atherosclerosis [Atherosclerotic heart disease] | Cardiovascular | 39,668 | 315,052 | 1.05 (1.04-1.06) | 1.10x10-16 |
| Atherosclerosis [ASCVD] | Cardiovascular | 45,807 | 305,469 | 1.04 (1.03-1.05) | 3.72x10-14 |
| Ischemic heart disease | Cardiovascular | 44,858 | 308,158 | 1.04 (1.03-1.05) | 4.03x10-14 |
| Angina pectoris | Cardiovascular | 14,293 | 344,985 | 1.05 (1.03-1.07) | 3.05x10-8 |
| Ischemic cardiomyopathy* | Cardiovascular | 3,080 | 363,185 | 1.10 (1.06-1.14) | 4.51x10-8 |
| Occlusion and stenosis of precerebral arteries | Cardiovascular | 8,198 | 352,946 | 1.06 (1.03-1.08) | 5.47x10-7 |
| Other cerebrovascular disease | Cardiovascular | 14,963 | 342,321 | 1.04 (1.03-1.06) | 5.48x10-7 |
| Hyperlipidemia | Endocrine/Metab | 143,368 | 196,251 | 1.15 (1.14-1.16) | 6.64x10-266 |
| Hypercholesterolemia | Endocrine/Metab | 79,851 | 265,403 | 1.12 (1.11-1.13) | 5.39x10-152 |
| Pure hypercholesterolemia | Endocrine/Metab | 45,995 | 302,961 | 1.13 (1.11-1.14) | 9.48x10-117 |
| Mixed hyperlipidemia | Endocrine/Metab | 45,183 | 308,966 | 1.08 (1.07-1.09) | 1.72x10-55 |
| Hyperglyceridemia | Endocrine/Metab | 50,899 | 301,143 | 1.07 (1.06-1.08) | 6.57x10-45 |
| Disorders of lipoprotein metabolism and other lipidemias | Endocrine/Metab | 4,184 | 358,110 | 1.09 (1.05-1.12) | 5.37x10-8 |
| Fatty liver disease (FLD) | Gastrointestinal | 18,234 | 337,190 | 0.94 (0.92-0.95) | 7.16x10-17 |
| Chronic liver disease | Gastrointestinal | 26,591 | 326,039 | 0.95 (0.94-0.96) | 1.91x10-14 |
| Chronic nonalcoholic liver disease | Gastrointestinal | 11,027 | 349,718 | 0.96 (0.94-0.98) | 1.25x10-5 |
| Familial hypercholesterolemia* | Genetic | 947 | 366,242 | 1.14 (1.08-1.21) | 1.16x10-5 |
| Neurocognitive disorder* | Mental | 167 | 367,393 | 1.38 (1.21-1.58) | 2.37x10-6 |
| Dementias | Neurological | 4,021 | 361,476 | 1.31 (1.28-1.35) | 9.93x10-78 |
| Alzheimer's disease | Neurological | 1,030 | 366,205 | 1.67 (1.58-1.77) | 9.33x10-75 |
| Dementias and cerebral degeneration | Neurological | 4,859 | 358,742 | 1.27 (1.24-1.30) | 7.04x10-71 |
| Memory loss | Neurological | 13,036 | 346,620 | 1.15 (1.13-1.17) | 1.71x10-58 |
| Symptoms and signs involving cognitive functions and awareness | Neurological | 28,556 | 322,629 | 1.08 (1.07-1.10) | 1.03x10-41 |
| Mild cognitive impairment | Neurological | 5,005 | 360,091 | 1.19 (1.16-1.23) | 3.80x10-40 |
| Vascular dementia | Neurological | 591 | 366,794 | 1.26 (1.17-1.36) | 9.44x10-10 |
| Altered mental status, unspecified | Neurological | 7,045 | 351,898 | 1.07 (1.04-1.09) | 2.43x10-8 |
| Dementia with Lewy bodies | Neurological | 138 | 367,537 | 1.42 (1.22-1.65) | 6.60x10-6 |

Bonferroni-significant phecodes (n = 27; α = 0.05 / 2,632 tested = 1.90 × 10<sup>−</sup>[illegible]) from the whole-cohort PheWAS (n = 367,757). Effect estimates come from logistic regression with the ordinal APOE genotype (ε2/ε2 < ε2/ε3 < ε3/ε3 < ε2/ε4 < ε3/ε4 < ε4/ε4) as the exposure, adjusted for age, sex at birth, EHR length, and the first 16 genetic principal components. Odds ratios (OR) are per rank increase in the ordinal APOE variable. Cases and controls reflect the analytic cohort after case-positive definition (≥2 phecode occurrences). Phecodes are grouped by category and sorted by ascending P value within category. Asterisks (\*) denote phecodes newly reaching Bonferroni significance in this analysis that were not tested (Ischemic cardiomyopathy, Familial hypercholesterolemia, Neurocognitive disorder) or not reported in the prior All of Us APOE PheWAS (Khajouei et al., 2025). PheTK package automatically excludes individuals with similar phenotypes from the control set for a given test, resulting in the number of cases + controls not matching exactly 367,757.

Comparing with the previous multi-ancestry PheWAS performed in All of Us ^11^, we identified 11 new significant associations ^11^ (Table 2). Nine phecodes were reported to be nominally significant in the previous study, such as vascular dementia (OR 1.26, 95% CI [1.17–1.36]; P = 9.44 × 10⁻¹ ), angina pectoris (OR 1.05, 95% CI [1.03–1.07]; P = 3.05 × 10⁻ ), chronic non-alcoholic liver disease (OR 0.96, 95% CI [0.94–0.98]; P = 1.25 × 10⁻ ) and familial hypercholesterolemia (OR 1.14, 95% CI [1.08–1.21]; P = 1.16 × 10⁻ ); two novel associations, dementia with Lewy bodies (OR 1.42, 95% CI [1.22–1.65]; P = 6.60 × 10⁻ ) and neurocognitive disorder (OR 1.38, 95% CI [1.21–1.58]; P = 2.37 × 10⁻ ) were not previously reported before due to insufficient case counts (< 50 cases) (Table 2). Sensitivity analysis was performed by including the following SDOH factors: education levels and SDI. All 27 associations remained significant after SDOH adjustment, confirming they are robust to potential confounders (Table S2).

To isolate ε2 and ε4-specific dosage effects beyond the ordinal model, we conducted additional analyses testing allele dosages of ε2 and ε4, each adjusted for copies of the other allele, using the same Bonferroni threshold as the primary PheWAS (p < 1.90×10⁻ ). The ε2 dosage yielded 13 significant associations (supplementary Figure 1 and Table S3). Most were protective effects on lipid and cardiovascular phecodes, and one neurological signal was detected for Alzheimer’s disease (OR = 0.62, P = 1.1×10⁻ ). The ε4 dosage yielded 22 significant associations, including risk effects for ten neurological phenotypes and nine lipid and cardiovascular phecodes (Supplementary Figure 2 and Table S4). ε4 dosage analysis also revealed an association with cerebral amyloid angiopathy (OR = 2.59, P = 1.2×10⁻ ) that was not detected in the primary ordinal PheWAS. Negative associations were observed for three gastrointestinal liver phecodes, consistent with the primary analysis. Together, these dosage analyses confirm that lipid effects are ε2/ε4-symmetric while neurological effects are driven predominantly by ε4. Interestingly, both ε2 and ε4 dose were positively associated with pure hyperglyceridemia.

To resolve whether APOE effects follow a linear dose-response or non-linear architecture, we performed diplotype analysis using ε3/ε3 as reference for the 27 significant hits, except for the phecode neurocognitive disorder, which no ε2/ε2 individuals were found in patients (Figure 2, Table S5). For lipidemia phecodes, effects were approximately linear, consistent with the ordinal model (Figure 2A). In contrast, dementia phecodes showed clear non-linearity, with ε4/ε4 conferring risks substantially larger than expected under linearity. The largest effects were observed for Alzheimer’s disease (OR = 7.73, 95% CI 6.07–9.86) and general dementias (OR = 3.45, 95% CI 2.96–4.01), followed by vascular dementia (OR = 2.80, 95% CI 1.88–4.18) and Lewy body dementia (OR = 3.85, 95% CI 1.66–8.94; n = 138). ε4/ε4 was also associated with increased risk of coronary atherosclerosis (OR = 1.16, 95% CI 1.07–1.25) and, notably, reduced risk of fatty liver disease (OR = 0.82, 95% CI 0.73–0.92). These patterns confirm the allele dosage effects identified and highlight the value of diplotype-resolved reporting for phenotypes with non-additive APOE risk architectures.

**Figure 2.**
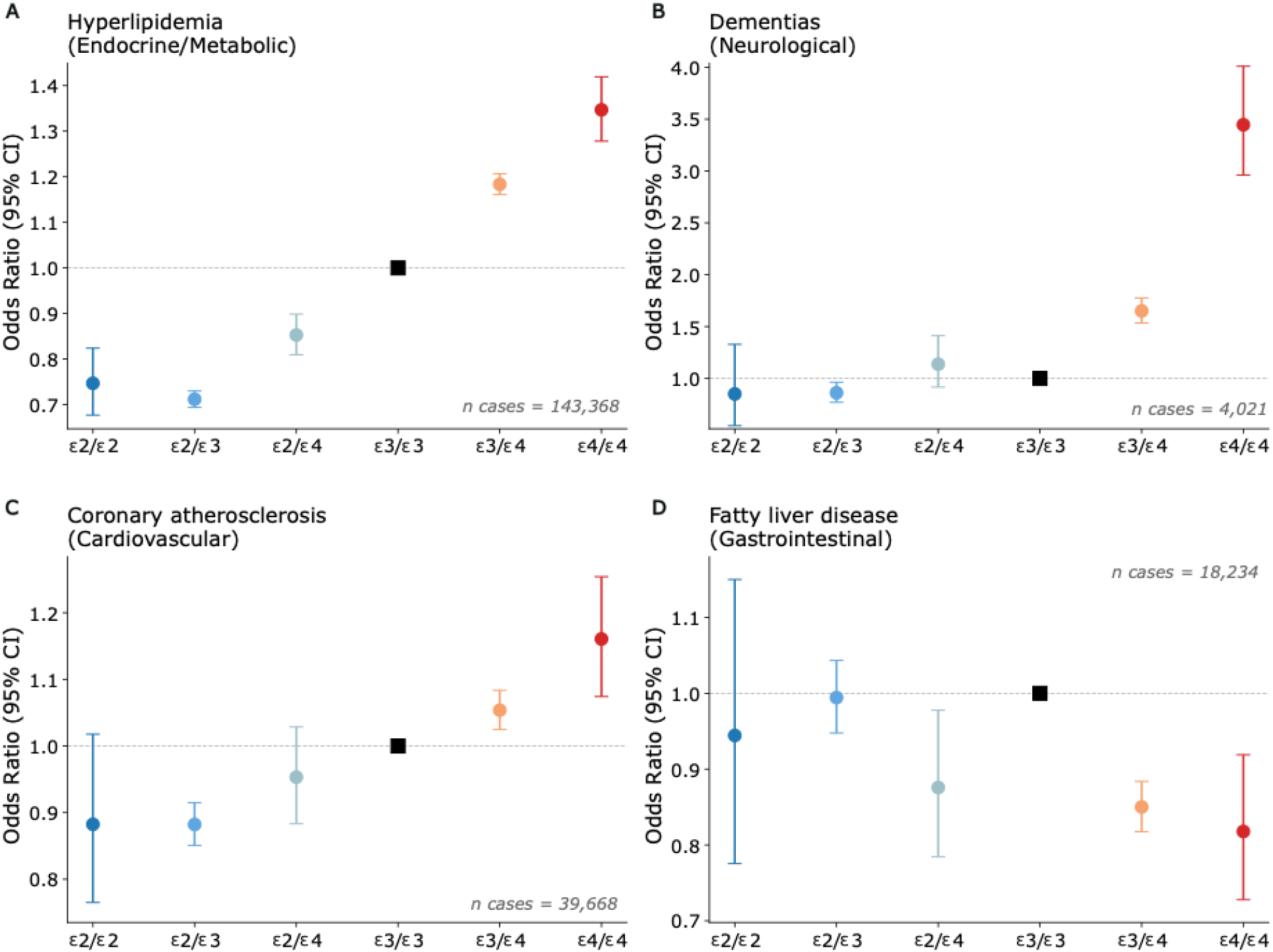
Disease risk by APOE diplotype. Forest plot displaying the risk of each diplotype for a) hyperlipidemia, b) dementias, c) coronary atherosclerosis and d) fatty liver disease. Each forest plot displays the 95% C.I for each genotype compared to ε3/ε3. Each logistic regression included age, sex, 16 genetic PCs and EHR length as covariates. We selected the top hit for the four largest PheWAS categories. Colors define each diplotype.

## Females carry higher APOE-related risk for lipid-related traits

To disentangle APOE sex-specific effects, we performed a stratified PheWAS in females (N = 224,916) and males (N = 142,841). In females, 2,384 phecodes were tested and 22 were found significant after Bonferroni correction (P < 2.10 × 10⁻ ) (Supplementary Figure 3 and Table S6); in males, 2,066 phecodes were tested and 19 were found significant (P < 2.42 × 10⁻ ) (Supplementary Figure 4 and Table S7). Among the 16 shared hits, we found the top overall-cohort associations, including Alzheimer’s disease, hyperlipidemia, and dementia. Female-only hits included familial hypercholesterolemia (OR 1.19, 95% CI [1.11–1.28]; P = 3.93 × 10⁻ ) and anoxic brain damage (OR 1.42, 95% CI [1.22–1.65]; P = 7.59 × 10⁻ ), among others. Vascular dementia was significant in males only (OR 1.28, 95% CI [1.16–1.41]; P = 1.66 × 10⁻ ).

To test whether APOE effects differ between sexes, we assessed heterogeneity across the 27 primary hits using a Cochran’s Q test. Five phecodes showed FDR-significant sex heterogeneity (FDR-P < 0.05, Figure 3, Table S8), all lipid-related and with stronger effects in females: hyperlipidemia, hypercholesterolemia, pure hypercholesterolemia, mixed hyperlipidemia, and hyperglyceridemia. No evidence of sex heterogeneity was observed for neurological phecodes, consistent with Khajouei et al. (Figure 3). Sensitivity analysis included testing for interaction between APOE and sex using an APOE×Sex product term in the logistic regression model. Results revealed a significant interaction for all five lipid phecodes after FDR correction: hyperlipidemia (FDR-P = 4.3×10⁻¹ ), hypercholesterolemia (FDR-P = 6.1×10⁻¹¹), pure hypercholesterolemia (FDR-P = 1.8×10⁻ ), mixed hyperlipidemia (FDR-P = 4.6×10⁻ ), and hyperglyceridemia (FDR-P = 1.8×10⁻ ). These results identify lipid-related phecodes (with approximately linear APOE dose-response) as the only ones with sex heterogeneity among the 27 primary hits, with no evidence of sex heterogeneity for neurological or cognitive phenotypes.

**Figure 3.**
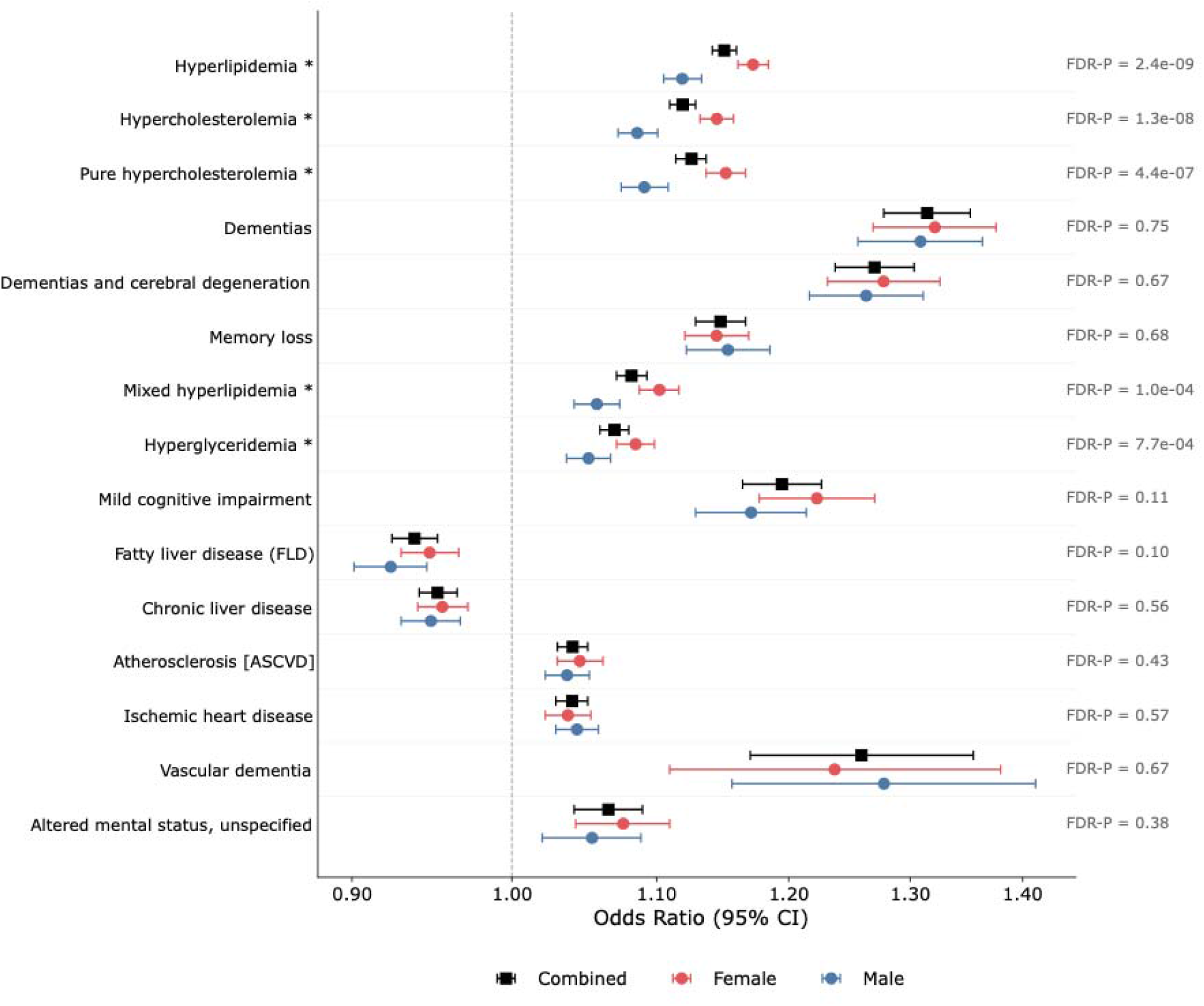
Sex-stratified analysis for APOE-associated phenotypes. Forest plot with Odds Ratios and 95% confidence intervals for male (blue), female (red) and combined (black). Logistic regressions for each phecode were adjusted by age, 16 genetic PCs and EHR length. Heterogeneity P-values were adjusted using the FDR method. Heterogeneity was only observed for lipid-related traits (asterisks). Only 15 out of 27 hits were displayed, including significantly heterogeneous phenotypes (5) and 10 non-significant ones, mainly for neurological, cardiovascular and liver.

## APOE-conferred risk varies across ancestries for hepatic, neurological and lipid phenotypes

To understand ancestry-specific effects of APOE alleles, we conducted an ancestry-stratified PheWAS for EUR, AFR, AMR, EAS, SAS, and MID, including 2,272, 1,824, 1,843, 901, 486, and 335 phecodes with at least 50 cases, respectively. EUR, AFR, AMR, EAS specific PheWAS revealed 23, 10, 6 and 2 significant associations after Bonferroni correction, with most significant phecodes being hyperlipidemia and hypercholesterolemia (Supplementary Figure 5-8, Table S9) in each ancestry.

No significant associations were found for SAS or MID likely due to insufficient statistical power. All ancestry-specific Bonferroni-significant phecodes were also identified in the overall cohort, except for Alcohol use disorder, which was only found in AMR.

Next, we performed ancestry-specific heterogeneity analyses for the 27 primary hits, excluding SAS and MID due to insufficient case counts. Nine phecodes showed significant heterogeneity across four ancestries (EUR, AFR, AMR, EAS) after FDR correction (Figure 4). Heterogeneity was maintained when excluding EAS which comprised of lowest sample size, confirming that heterogeneity is not driven by lack of power (Table S10). Three of these replicated ancestry-heterogeneous associations were previously reported ^11^: hyperlipidemia, mild cognitive impairment, and symptoms and signs involving cognitive functions and awareness. The remaining six were novel ancestry-heterogeneous findings: hypercholesterolemia, dementias, dementias and cerebral degeneration, memory loss, chronic liver disease, and fatty liver disease. For dementia-related phecodes, the largest effect size was found in EUR, with attenuation in the other three groups. For hyperlipidemia, the largest effect size was found in EAS. Interestingly, APOE did not show protective effects for fatty liver disease and chronic liver disease in AFR, contrary to other ancestry groups (Figure 4). These findings confirm ancestry heterogeneity across three categories of the primary PheWAS: endocrine/metabolic, neurological, and gastrointestinal, with the largest lipid effect in EAS and the largest dementia effect in EUR. We did not observe heterogeneity in cardiovascular phenotypes.

**Figure 4.**
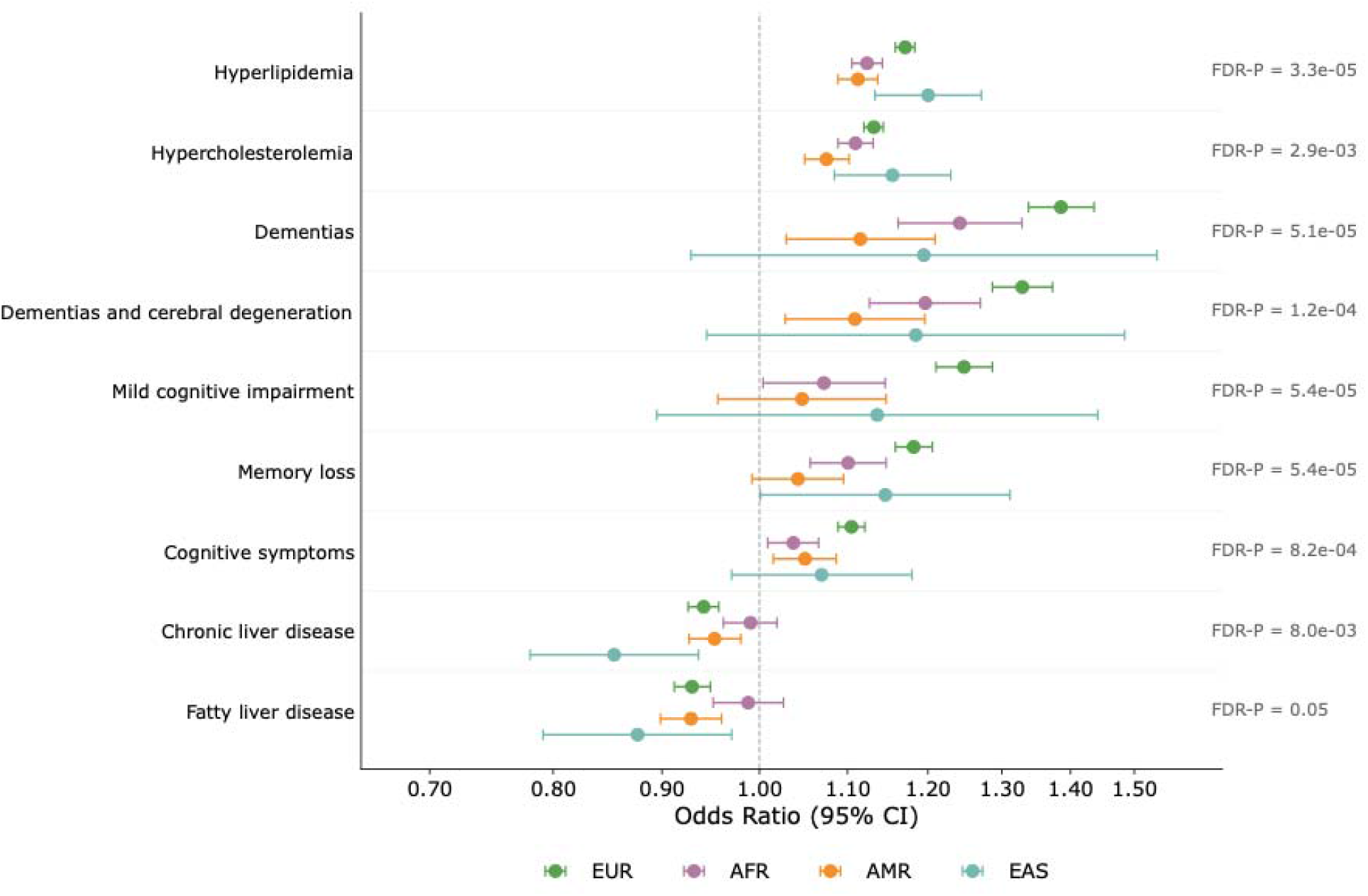
Ancestry-stratified risk for APOE-associated phenotypes. Forest plot displaying nine phenotypes that demonstrated heterogeneity between four ancestry groups: EUR, AFR, AMR and EAS. The x-axis shows the Odds Ratio for the APOE ordinal model and its respective 95% Confidence interval. Logistic regressions for each phecode were adjusted by age, sex, 16 genetic PCs and EHR length. Heterogeneity P-values were adjusted using the FDR method.

## Proteomic signatures of APOE genotypes across ancestries

To identify proteomic signatures associated with APOE allele dosage, we performed differential protein level analysis using ε2 and ε4 dosage in linear regression models adjusted for age, sex, 16 genetic principal components, and plate ID. As expected, we found a positive association between APOE level and ε2 dosage (β = +0.874) and a negative association with ε4 (β =-0.846), consistent with previous reports ^17^. Additionally, ε4 dosage was associated with 17 proteins (Figure 5A and Table S11) and ε2 dosage (Figure 5B and Table S12) with 77 proteins at FDR < 0.05. Top ε4-associated proteins included MENT, SNAP25, PALM, and CSNK2A1; top ε2-associated proteins included LDLR, MENT, PLA2G7, and BRK1. These proteins were involved in pathways of lipid metabolism (APOA1, APOD, LDLR), neurodegeneration (SNAP25, PALM, CSNK2A1), and immune regulation.

**Figure 5.**
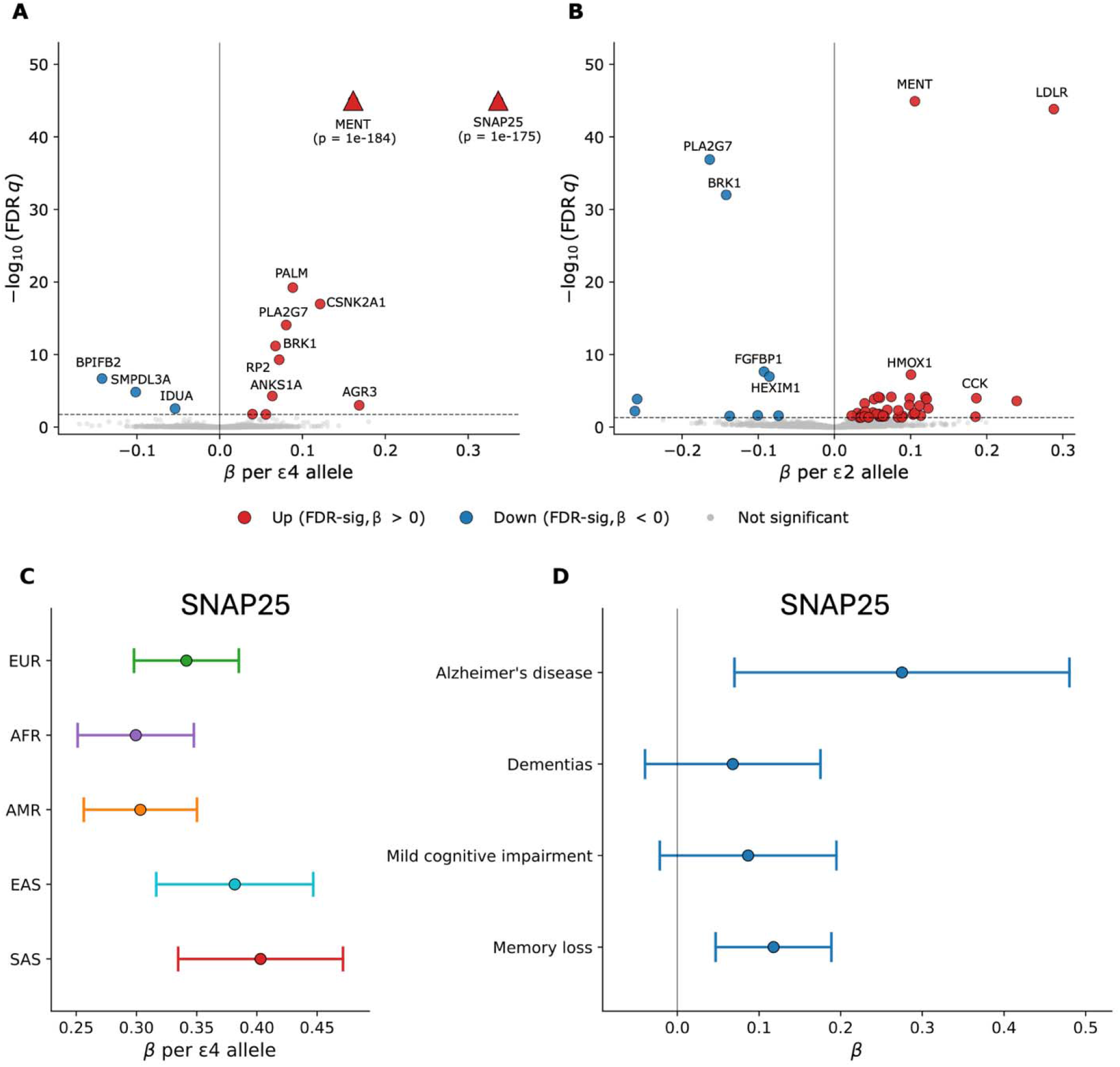
APOE. ε**2 and** ε**4 proteomics signatures across ancestries.** Volcano plots showing proteins associated with ε4 allele dosage (A) and ε2 dosage (B). Significantly upregulated proteins are colored in red, and significantly downregulated proteins in blue. The X-axis shows beta per e4 allele (A) and per e2 allele (B), and the Y-axis displays the -log10(P-value) after FDR correction. We capped the capped -log10(FDR p) at 45, but the real value is displayed for capped proteins (triangles).

Our proteomic findings both validated and extended prior European-ancestry work. Consistent with Lu et al. ^17^, we recovered several APOE-associated proteins on both SomaLogic and Olink platforms, including LDLR, PLA2G7, PLA2G10, ANGPTL3, HMOX1, MENT, and BRK1, spanning lipid metabolism, immune regulation, and previously reported APOE signatures. Beyond these, the multi-ancestry design and expanded Olink Explore HT coverage enabled discovery of proteins not previously reported, including SNAP25, PALM, CSNK2A1, and AGR3 for ε4, and LPA, CCK, APOA1, and APOD for ε2. Notably, we identified a novel protein, SNAP25, which was not reported in the previous proteomics study ^17^.

We next tested for ancestry specific APOE-proteomic effects. We repeated the joint ε2/ε4 model stratified by genetic ancestry across five major groups (EUR n=2,648; AFR n=1,439; AMR n=2,033; EAS n=1,374; SAS n=1,187). For ε2 dosage, we identified 8, 6, 4, 4, and 2 FDR-significant proteins in EUR, AFR, AMR, EAS, and SAS, respectively (Table S13) of which five exhibited significant ancestry heterogeneity. APOA1 showed a positive association with ε2 dosage that was strongest in EUR (β = +0.204, P = 3.3×10⁻ ) and AMR (β = +0.181, P = 0.011), attenuated in AFR (β = +0.106, P = 0.11), and absent in EAS (β = −0.061, P = 0.34) and SAS (β = −0.048, P = 0.57), indicating that the ε2-mediated increase in APOA1 is largely restricted to non-Asian populations (Q FDR-P = 0.02). In contrast, ε4 dosage was uniformly associated with lower APOA1 across all five ancestries (β range −0.04 to −0.13), with nominal significance in EUR, EAS, and SAS, consistent with ε4’s established effect on HDL apolipoprotein reduction. Additional ancestry-specific ε2 findings included CDC42EP1 (β = −0.18, FDR-P = 9.3×10⁻³), PDGFC (β=+0.26, FDR-P = 1.05×10^-2^) in AFR, FGFBP1 (β=−0.35, FDR-P=1.39×10⁻ ), LPA (β=−0.28, FDR-P = 5.51×10⁻³) in EAS, and ITGB5 in AMR (β=+0.28, FDR-P = 2.47×10⁻²), the latter implicated in extracellular matrix organization and neurodegeneration.

For ε4 dosage, we identified 13, 5, 3, 3, and 3 FDR-significant proteins in EUR, AFR, AMR, EAS, and SAS, respectively (Table S14). Two proteins were found significant only in AFR: PLA2G7 (β=+0.23, FDR-P = 6.34×10⁻ ), IL32 (β=+0.19, FDR-P = 3.33×10⁻²).

Five proteins exhibited significant ancestry heterogeneity by Cochran’s Q test (FDR-P < 0.05; Figure 6). three were EUR-specific (BMERB1, β = +0.185; SMPD1, β = −0.191; SMPDL3A, β = −0.184). On the other hand, SNAP25 showed consistent effect sizes across ancestries (Figure 5C). Further analysis showed that SNAP25 levels are higher in Alzheimer’s disease (β = +0.476, FDR-P = 1.5×10⁻²) and memory loss cases (β = +0.200, FDR-P = 4.69×10⁻³) compared to controls (Figure 5D and Table S15), supporting its role as a neurodegeneration biomarker.

**Figure 6.**
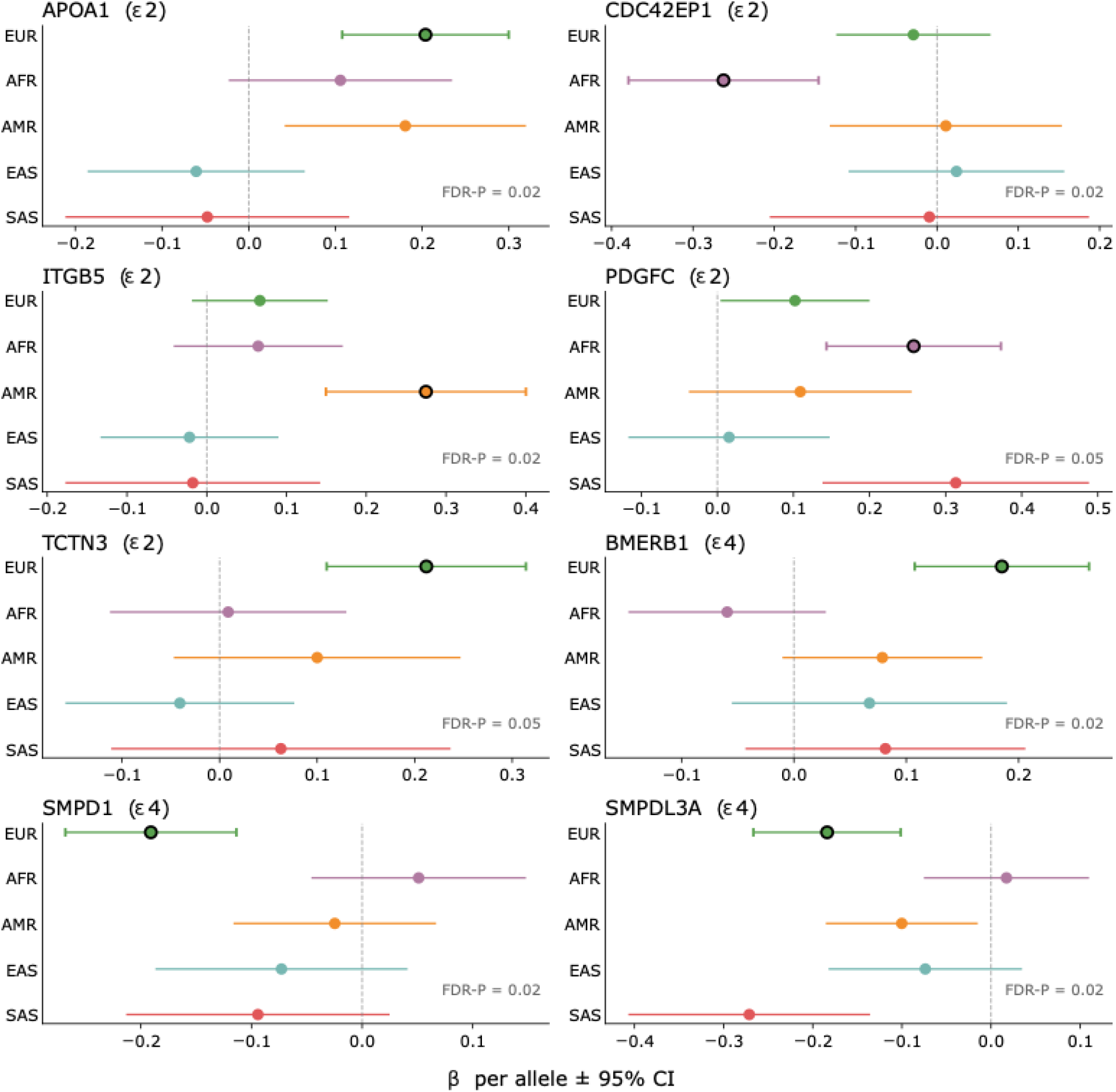
APOE. ε**2 and** ε**4 associated proteins with heterogeneous effects across ancestries.** Forest plot with 8 proteins that showed ancestry-heterogeneity in their associations with either ε2 or ε4 allele dosage. FDR corrected P-value from the heterogeneity test are displayed for each forest plot. The ancestry where the protein was found significant has a dark circle. X-axis shows the effect size and 95% confidence interval. Y-axis shows the ancestries included (EUR, AFR, AMR, EAS and SAS).

## Discussion

To our knowledge, this is the largest multi-ancestry APOE PheWAS to date, combining a PheWAS of 367,757 participants with the first ancestry-stratified plasma proteomic analysis that includes AMR and EAS ancestries. We identified 27 phenotypes associated with APOE — 11 not previously reported in this cohort — converging on four categories: dyslipidemia (Endocrine/Metabolic), dementia and cognitive decline (Neurological), coronary atherosclerosis (Cardiovascular), and protection against hepatic lipid accumulation (Gastrointestinal). Across these themes, five lipid-related phecodes showed sex specific effects and nine phecodes for ancestry specific effects, with the largest lipid and hepatic effects in EAS and the largest dementia effect in EUR. Plasma proteomics analysis identified SNAP25 as an ancestry-portable candidate biomarker of APOE-ε4 not previously detectable at scale, alongside ancestry-specific proteins (IL32 in AFR, ITGB5 in AMR, APOA1 in non-Asian populations) that may provide molecular hypotheses for the observed phenotypic heterogeneity. Together, these findings strongly supported that genetic ancestry should be integrated into APOE-based risk interpretation.

As expected, we replicated 16 of the 17 top phecode associations reported by Khajouei et al. in an expanded cohort of 367,757 All of Us participants, approximately double the previous sample size (n = 181,880) ^11^. Non-inflammatory disorders of vulva and perineum, which was reported in Khajouei et al. (P = 1.07 × 10⁻ ), did not reach Bonferroni significance in our sex-adjusted overall-cohort analysis, though the direction of effect was preserved. New associations included vascular dementia, angina pectoris, unstable angina, occlusion/stenosis of precerebral arteries, highlighting the role of APOE in cardiovascular and neurological phenotypes beyond classic dementias and hyperlipidemia, and the increased power compared to previous studies ^11^.

We found a significant association with vascular dementia that was not reported before using All of Us due to lack of statistical power. Our diplotype analysis confirmed these findings, with an OR of 1.55 (95% C.I = 1.29 – 1.88) for ε3/ε4 and 2.8 (95% C.I = 1.88 – 4.18) for ε4/ε4. Associations between APOE ε4 and vascular dementia have been documented before, with OR = 1.65 (95% CI = 1.40–1.94) for ε3/ε4 and OR = 3.17 (95% CI = 2.09–4.80) for ε4/ε4 ^21^. This finding along with precerebral artery occlusion/stenosis suggest that the APOE role on the vasculature is consistent across ancestries (no heterogeneity observed), although we might lack power to detect differences between ancestries and sexes. APOE ε4 has been shown to affect the vasculature through several mechanisms, including blood brain barrier dysfunction ^22^, amyloid beta plaques clearance impairment ^23^, extracellular matrix remodeling, vascular metabolism ^24^, among others. Our findings confirm well-established associations from the literature with similar estimates, confirming our increased power to detect associations that were not possible before the last release from All of Us ^11^.

We also confirmed the recently reported protective association of APOE ε4 with fatty liver disease and chronic liver disease. This protection showed significant ancestry heterogeneity for chronic liver disease (Cochran’s Q FDR-P = 8×10⁻³) and fatty liver disease (Cochran’s Q FDR-P = 0.05). The reduced risk was consistent in EUR and EAS but absent in AFR, which could be due to African populations have substantially greater local diversity around APOE, which resulted in attenuated hepatic effect in Africans. The opposing direction of the ε4 effect on circulating lipid disorders (increased risk of hyperlipidemia and hypercholesterolemia) versus hepatic fat is consistent with a lipid-partitioning model, in which APOE ε4 favors lipid retention in the circulation and peripheral adipose over hepatic storage in mouse models ^25^. These results demonstrate the interplay between risk and protection given by the APOE alleles, highlighting the complexity given the metabolic processes in which APOE is involved.

Ancestry heterogeneity was also found in phenotypes where APOE genotypes do not show a linear relationship such as dementia (Cochran’s Q FDR-P = 5.1 x10^-5^), memory loss (Cochran’s Q FDR-P = 5.4x10^-5^), and mild cognitive impairment (Cochran’s Q FDR-P = 5.4x10^-5^). EUR shows a larger risk of these neurological phenotypes, followed by AFR and AMR. Recent reports have shown that APOE has heterogeneous effects in Latinos, including smaller risks of AD associated with APOE ε4 compared to Europeans^26^. Although several reasons have been proposed to explain these differences, including local ancestry, confounding by admixture, among others, the exact cause remains an open question. In AFR, a recent common 19bp deletion in the APOE enhancer region protects against AD and interacts with APOE genotypes ^27^. We tested the most common confounders, including SDOH, and heterogeneity remained, suggesting that other factors such as interaction with other variants (epistasis) or environmental factors could contribute to this difference.

Sex heterogeneity was found exclusively in lipid-related phenotypes, with no evidence of heterogeneity for neurological outcomes. This association is consistent with hormonally mediated regulation of APOE: estrogen upregulates APOE expression, providing lipid protection before menopause ^28,29^ and loss of this protection after menopause exacerbates the APOE-ε4–driven downregulation of LDLR ^17^, yielding disproportionate lipid-related risk in women ^30–32^, consistent with our observation of stronger effects in female carriers for hyperlipidemia, hypercholesterolemia, and mixed hyperlipidemia.

The lack of sex heterogeneity for neurological phenotypes may reflect the non-linear architecture of APOE-associated dementia risk. For instance, several reports have suggested sex-specific ε4 effects on cognitive decline ^33–36^. Main difference between our study and the ones that reported a higher risk in female is that they either evaluated this association longitudinally (with proportional hazard models) and they measured cognitive decline in multiple visits ^33–36^, therefore providing stronger evidence that our cross-sectional approach.

Finally, proteomic analysis revealed proteins robustly associated with APOE allele dosage across ancestries such as SNAP25. SNAP25 plays an essential role in the exocytosis of neurotransmitters, as it helps in the fusion with the cell membrane ^37^. SNAP25 has been proposed as a biomarker of synaptic loss and brain injury ^38^. This evidence is concordant with the higher levels of SNAP25 found in Alzheimer’s disease and memory loss cases (Figure 5D). A few studies have identified SNAP25 as an APOE and AD biomarker in CSF ^17,39^. However, the largest APOE proteomics study did not identify it as an APOE-associated protein in plasma (β = 0.008, P = 0.74) ^17^. This discrepancy was due to platform discordance between SomaLogic and Olink, where abovementioned study used SomaScan panel to quantify proteins. It was highly likely originated from SomaScan assay specificity: that altered aptamer binding was driven by SERPINA1 proteoforms, as strong SNAP25 cis-pQTL was only identified in Olink panel (rs362562), while only exceptionally strong SERPINA1-region trans-pQTL was found in SomaLogic related proteomic GWAS for SNAP25 ^40,41^. In addition, studies have shown that the correlation of protein levels between CSF and plasma is generally low, especially for neurological phenotypes ^42,43^. Given the non-invasive nature of a blood draw, SNAP25 could be a novel biomarker of APOE ε4 effects across ancestries, and it could be used to track memory loss and AD, as recently suggested by preliminary studies and our results (Figure 5D) ^44,45^.

APOA1 showed significant ancestry heterogeneity in the association with ε2 dosage: no significant association in EAS and SAS populations but positive association in EUR and AMR. APOA1 levels are positively associated with HDL levels, which takes excess cholesterol in the bloodstream and delivers it to the liver, known as reverse transport ^46^. The absence of an ε2-driven APOA1 response in EAS could explain the larger effects of APOE genotype (ordinal) on EAS for lipid-related traits, and the largest protective effects against fatty liver disease, as reverse cholesterol transport might be diminished due to lack of APOA1.

Four proteins were significant only in AFR: CDC42EP1, PDGFC (ε2 dosage), IL32 and PLA2G7 (ε4 dosage) (Table S13 and 14). CDC42EP1 is a CDC42 binding protein that mediates actin cytoskeleton reorganization ^47^. CDC42 regulates neurite outgrowth, neurotransmitter and T cell mediated neuroinflammation ^48^. Moreover, CDC42 pathway activity has been linked with cognitive impairment, neuron senescence and synaptic loss ^48,49^. Downregulation of CDC42EP1 could help reduce inflammation, possibly explaining one of the protective mechanisms of APOE ε2 in AFR, and the smaller risk observed compared to EUR and EAS for lipid-related and neurological traits.

In contrast, IL32 was associated with APOE ε4 alleles only in AFR. IL32 is associated with chronic inflammation, endothelial function and HDL concentrations ^50^. IL32 is linked to cardiovascular diseases and atherosclerosis ^51,52^. Moreover, IL32 has been associated with neuroinflammation, memory impairment, glia activation and amyloidogenesis ^53,54^. These associations are consistent with the observed effects of APOE ε4 on lipid-related traits and neurological phenotypes. Inflammation might be different between ancestry groups ^55^, influencing responses to microorganisms ^56^. IL32 has different isoforms, and genetic variation that is ancestry-specific could contribute to the differences observed in APOE ε4 related signatures ^57^. In summary, IL32 could be mediating some of the effects of APOE ε4 in the AFR population, but further validation in independent cohorts is needed.

We would like to acknowledge several limitations in this study. Reduced sample sizes for EAS, SAS and MID have limited statistical power to detect heterogeneity between ancestries, especially for neurological phenotypes. The use of EHR might result in reduced diagnostic accuracy, although we used a strict definition with at least 2 code occurrences per phecode to be classified as a case. This bias due to EHR case definition could lead to underestimated ORs, particularly if cases were misclassified or if we included controls with subclinical AD or another phenotype. In addition, dyslipidemias were classified using phecodes, and we cannot ascertain if blood measurements were used to perform these diagnoses. Similarly, biases due to geographical variables could result in differences in billing and cognitive assessment.

SDOH was included as a covariate in our analyses to help mitigate confounding from these sources, however, it might not be sufficient to account for all social-economical differences. Finally, even with increased sample size in our study, the number of ε2/ε2 carriers were still low (N=2,376), particularly for less common diseases such as neurodegenerative disorders which are generally underrepresented in population-based biobanks.

In summary, our findings provide robust evidence to support the need to ancestry integrated APOE-risk interpretation. Across 367,757 participants of six ancestries, the risks and protections conferred by APOE differ meaningfully between populations — for hepatic, dementia, and lipid phenotypes — and the plasma proteins we identify (SNAP25 shared across ancestries; IL32 in AFR, ITGB5 in AMR, APOA1 in non-Asian populations) parallel these differences and suggest biomarkers for ancestry-specific follow-up. Prospective evaluation of ancestry-aware APOE risk models and validation of these plasma proteomic candidates, including SNAP25 as a candidate cross-ancestry ε4 biomarker, are the next steps toward translating APOE genetics into equitable clinical practice.

## Contributors

A.M-G and S.Z conceived and designed the study. A.M-G performed the genetic ancestry and APOE genotype extraction, phecode ascertainment, PheWAS analyses, proteomic analyses, and drafted the manuscript. A.M-G, S.Z, T.M.Z, J.J, Z.W, H.M.T, P.V-C, C-Y.S and C.A.O contributed to the statistical analysis and interpretation. A.M-G, T.M.Z, C-Y.S and Z.W contributed to the proteomic data processing. A.M-G, H.M.T, T.M.Z and Z.W accessed and verified the underlying data. S.Z supervised the study. All authors reviewed, edited, and approved the final manuscript, and had full access to all data in the study and accept responsibility for the decision to submit for publication.

## Data Availability

Individual-level data from the All of Us Research Program are not publicly available but can be accessed by approved researchers through the All of Us Researcher Workbench (https://workbench.researchallofus.org) upon completion of the required training and institutional data use agreement. All analytical code used in this study will be available in the All of Us workbench. Summary statistics for the PheWAS and proteomic analyses are provided in the Supplementary material. No individual participant data will be shared outside the All of Us Researcher Workbench in accordance with the program's data access policy.

## Acknowledgments

We would like to thank All of Us participants for their contribution to the research program. A.M-G is supported by STAGE Quebec and the Canadian Institute for Health Research (CIHR) doctoral scholarship, Funding reference number: 210230. S.Z. is supported by a Fonds de recherche du Québec santé (FRQS) Chercheurs-boursiers J1 award. This research was supported by the Canadian Institute for Health Research (CIHR) project grant PJT205932 (S.Z).

