## Supplementary information for "Phenome-wide and proteomic analysis of APOE alleles across different ancestries in the All of Us Research Program"

**Short title:** APOE PheWAS and proteomic analysis in All of Us

### SUPPLEMENTARY METHODS

#### Study cohort

The All of Us research program is a multi-ancestry cohort from the U.S that recruited individuals aged at least 18 years, and it hosts whole-genome sequencing, proteomics and electronic health records (EHR) data. We first retrieved genotype data for 535,000 individuals using the exome data from WGS, as both variants used to define APOE genotypes are in exon 4 of the APOE gene. 28 individuals have a missing APOE genotype due to missingness in either one of the variants needed for assignment. After assigning APOE genotype using a Python script that uses genotypes for each variant in PLINK format as input, we constructed a cohort using the PheTK Python package. To add covariates of interest (EHR length, genetic PCs, age and Sex), we use the add.covariates function in the PheTK package, which uses the person ID as input to find the corresponding covariates. We removed individuals with any missing data in terms of covariates, keeping a final cohort with 367,757 individuals. Finally, we used the PheTK to perform the PheWAS, requiring at least 2 recorded phecodes for every case and at least 50 cases per phenotype to be included in the analysis.

#### APOE genotype and allele dosage

APOE diplotypes were derived from two single-nucleotide polymorphisms, rs429358 and rs7412, extracted from the Whole-genome sequencing, restricted to coding variants (Exome subset) available in the All of Us Researcher Workbench, as described previously (1). From 535,662 individuals with WGS data, 181 has missing genotypes for at least of the SNPs needed to define APOE diplotyes. The combination of genotypes at these two variants determines the six common APOE diplotypes (ε2/ε2, ε2/ε3, ε2/ε4, ε3/ε3, ε3/ε4, and ε4/ε4). Individuals whose diplotype could not be unambiguously assigned were excluded (N=181). From the diplotype, we additionally derived two continuous variables representing allele dosages: copies_ε2 (0, 1, or 2) and copies_ε4 (0, 1, or 2), corresponding to the number of ε2 and ε4 alleles carried by each participant.

#### Primary statistical analysis

The primary phenome-wide association study (PheWAS) treated APOE as an ordinal variable (APOE_ord) reflecting the previously published Alzheimer's disease risk hierarchy of ε2/ε2 < ε2/ε3 < ε3/ε3 < ε2/ε4 < ε3/ε4 < ε4/ε4, coded as integers 0 through 5. Association testing was performed using logistic regression via the PheTK package, with each phecode modeled independently as a binary outcome. All models were adjusted for age at analysis, sex assigned at birth, EHR length, and the first 16 genetic PCs. A Bonferroni-corrected significance threshold of $\alpha = 1.90\times10^{-5}$ (0.05 / 2,632 phecodes) was applied.

### Analysis adjusting for Social Determinants of Health

To assess sensitivity to potential ascertainment and detection biases correlated with socioeconomic factors, we conducted a Social Determinants of Health (SDOH)-adjusted PheWAS. SDOH data were obtained from the All of Us "zip_code_socioeconomic" table, which provides individual-level information linked to participants' three-digit ZIP Code Tabulation Areas. Two SDOH covariates were included: highest attained education level, dichotomized as college degree or higher versus less than college, and the raw Social Deprivation Index (SDI). Household income was collected but had substantial missingness in the analytical cohort and was excluded from adjustment. Individuals with missing SDOH data were removed, resulting in an analytical cohort of 345,995 participants and 2,605 tested phecodes. All other model specifications matched the primary analysis. A Bonferroni threshold of $\alpha = 1.92\times10^{-5}$ (0.05 / 2,605 phecodes) was applied.

### Sex-stratified analysis and APOE by Sex interaction

To evaluate whether APOE associations differed between female and male participants, sex-stratified PheWAS were conducted using the same modeling approach and covariate set as the primary analysis (excluding sex assigned at birth). A total of 2,385 phecodes were tested in females (2,384 converged) and 2,067 phecodes in males (2,066 converged). Effect estimates for phecodes reaching Bonferroni significance in the primary analysis (n = 27) were compared across sex strata using a Cochran's Q test. Benjamini-Hochberg false discovery rate (FDR) correction was applied across the 27 tested phecodes, with a threshold of $p < 0.05$.

For phecodes exhibiting FDR-significant sex heterogeneity, we additionally fit pooled logistic regression models incorporating an APOE_ord × sex_at_birth interaction term. Interaction models were adjusted for age, sex assigned at birth, EHR length, and the first 16 PCs, and were fit using the Python statsmodels library v.0.14.6. Interaction p-values were subjected to FDR correction across the five tested phecodes.

### Diplotype-specific analysis

To characterize APOE associations without imposing an additivity or ordinal-ranking assumption, we performed logistic regression using categorical diplotype dummies for the top Bonferroni-significant hits from the primary analysis (n = 27). Each of the five non-reference diplotypes (ε2/ε2, ε2/ε3, ε2/ε4, ε3/ε4, ε4/ε4) was contrasted against ε3/ε3 as the reference group. Models were adjusted for age, sex assigned at birth, EHR length, and the first 16 PCs. Case/control status was defined identically to the primary PheWAS (≥2 phecode instances for cases; zero for controls). Analyses were performed using the Python statsmodels library.

### Allele dosage sensitivity analysis

To evaluate robustness of our ordinal approach, we conducted additional phenome-wide analyses using allele dosages of ε2 and ε4 as the independent variable of interest. Two models were tested: (i) copies_ε4 with copies_ε2 included as a covariate; and (ii) copies_ε2 with copies_ε4 included as a covariate. These models aim to isolate each allele's independent effect from the mechanical anti-correlation between allele counts at the APOE locus (copies_ε2 + copies_ε3 + copies_ε4 = 2). All analyses were adjusted for age, sex assigned at birth, EHR length, and the first 16 PCs. Analyses were performed with the PheTK package, and Bonferroni correction was applied at $\alpha = 1.90\times10^{-5}$ (0.05 / 2,632 phecodes).

#### Proteomics analysis

Proteomic data from Olink was retrieved from the All of Us Workbench, which includes 5,416 proteins for 9,841 participants. We selected batch-normalized data without replicates for subsequent analysis. Details about the quality control performed by All of Us can be found here: https://support.researchallofus.org/hc/en-us/articles/50655639562900-All-of-Us-Genomics-Multi-omics-Quality-Report. NPX values were retrieved and use as the abundance metric for every protein measured. As a quality control step, we filtered out individuals with > 15% of missing proteins (190 individuals). In addition, we removed protein values that were outside 5 standard deviations in the whole cohort. Finally, we applied inverse rank normalization to make sure that protein values follow a normal distribution. For all analyses, we adapted the allele dosage model, including both e2 and 4 dosages as independent variables in the model, adjusting by 16 genetic PCs, sex, age at sample collection and plate ID (to account for technical variation). The final cohort with all covariates included 9,132 individuals and 5,415 proteins (we excluded APOE). We included APOE as a positive control, as we know the variants used to define APOE genotypes affect protein abundance, as previously reported (2).

### Supplementary Tables

**Table S1.** Definition of APOE genotypes

| *APOE* Genotype | rs429358 | rs7412 |
|---|---|---|
| ε1ε1 | CC | TT |
| ε1ε2 | CT | TT |
| ε2ε4 | CT | CT |
| ε1ε4 | CC | CT |
| ε2ε2 | TT | TT |
| ε2ε3 | TT | CT |
| ε3ε3 | TT | CC |
| ε3ε4 | CT | CC |
| ε4ε4 | CC | CC |

**Table S2.** APOE associations for the 27 significant whole-cohort PheWAS phecodes after adjustment for social determinants of health.

| Phecode | Phenotype | Category | Cases / Controls | OR (95% CI) | β (SE) | P |
|---|---|---|---|---|---|---|
| EM_239 | Hyperlipidemia | Endocrine/Metab | 135,794 / 183,647 | 1.16 (1.15–1.16) | +0.144 (0.004) | 1.80e-264 |
| EM_239.1 | Hypercholesterolemia | Endocrine/Metab | 76,018 / 248,738 | 1.12 (1.12–1.13) | +0.117 (0.004) | 3.24e-156 |
| EM_239.11 | Pure hypercholesterolemia | Endocrine/Metab | 43,945 / 284,241 | 1.13 (1.12–1.14) | +0.122 (0.005) | 4.86e-118 |
| NS_328.1 | Dementias | Neurological | 3,770 / 340,105 | 1.32 (1.28–1.36) | +0.276 (0.015) | 4.90e-74 |
| NS_328.11 | Alzheimer's disease | Neurological | 986 / 344,513 | 1.67 (1.58–1.77) | +0.515 (0.029) | 9.35e-72 |
| NS_328 | Dementias and cerebral degeneration | Neurological | 4,555 / 337,572 | 1.27 (1.24–1.31) | +0.240 (0.014) | 7.22e-67 |
| NS_329.1 | Memory loss | Neurological | 12,349 / 326,044 | 1.15 (1.13–1.17) | +0.140 (0.009) | 5.93e-57 |
| EM_239.3 | Mixed hyperlipidemia | Endocrine/Metab | 42,924 / 290,199 | 1.09 (1.08–1.10) | +0.083 (0.005) | 8.15e-58 |
| EM_239.2 | Hyperglyceridemia | Endocrine/Metab | 48,304 / 282,844 | 1.07 (1.06–1.08) | +0.072 (0.005) | 1.26e-47 |
| NS_329 | Symptoms and signs involving cognitive functions and awareness | Neurological | 26,703 / 303,954 | 1.09 (1.07–1.10) | +0.083 (0.006) | 1.22e-40 |
| NS_329.5 | Mild cognitive impairment | Neurological | 4,731 / 338,764 | 1.19 (1.16–1.23) | +0.175 (0.014) | 7.04e-37 |
| GI_542.2 | Fatty liver disease (FLD) | Gastrointestinal | 16,930 / 317,750 | 0.94 (0.92–0.95) | -0.067 (0.008) | 8.92e-17 |
| CV_404.2 | Coronary atherosclerosis [Atherosclerotic heart disease] | Cardiovascular | 37,356 / 296,395 | 1.05 (1.03–1.06) | +0.045 (0.006) | 2.49e-15 |
| GI_542 | Chronic liver disease | Gastrointestinal | 24,532 / 307,538 | 0.95 (0.94–0.96) | -0.053 (0.007) | 1.63e-15 |
| CV_436 | Atherosclerosis [ASCVD] | Cardiovascular | 43,124 / 287,378 | 1.04 (1.03–1.05) | +0.040 (0.005) | 2.70e-13 |
| CV_404 | Ischemic heart disease | Cardiovascular | 42,124 / 290,066 | 1.04 (1.03–1.05) | +0.039 (0.005) | 9.38e-13 |
| NS_328.12 | Vascular dementia | Neurological | 547 / 345,106 | 1.24 (1.15–1.34) | +0.216 (0.039) | 4.07e-08 |
| NS_329.8 | Altered mental status, unspecified | Neurological | 6,439 / 331,555 | 1.06 (1.04–1.09) | +0.063 (0.012) | 1.52e-07 |

| Phecode | Phenotype | Category | Cases / Controls | OR (95% CI) | β (SE) | P |
|---|---|---|---|---|---|---|
| CV_403 | Angina pectoris | Cardiovascular | 13,451 / 324,587 | 1.05 (1.03–1.07) | +0.047 (0.009) | 7.76e-08 |
| CV_414.5 | Ischemic cardiomyopathy* | Cardiovascular | 2,878 / 341,720 | 1.10 (1.06–1.14) | +0.095 (0.018) | 9.47e-08 |
| EM_244 | Disorders of lipoprotein metabolism and other lipidemias | Endocrine/Metab | 4,008 / 336,734 | 1.09 (1.05–1.12) | +0.082 (0.015) | 1.05e-07 |
| CV_433.2 | Occlusion and stenosis of precerebral arteries | Cardiovascular | 7,756 / 331,981 | 1.06 (1.04–1.08) | +0.057 (0.011) | 5.08e-07 |
| CV_433 | Other cerebrovascular disease | Cardiovascular | 14,085 / 321,997 | 1.04 (1.03–1.06) | +0.043 (0.008) | 3.69e-07 |
| MB_308.4 | Neurocognitive disorder* | Mental | 152 / 345,657 | 1.38 (1.20–1.59) | +0.324 (0.072) | 6.43e-06 |
| NS_328.14 | Dementia with Lewy bodies | Neurological | 131 / 345,787 | 1.41 (1.21–1.65) | +0.345 (0.079) | 1.39e-05 |
| GE_965.4 | Familial hypercholesterolemia* | Genetic | 899 / 344,550 | 1.14 (1.08–1.22) | +0.135 (0.031) | 1.59e-05 |
| GI_542.7 | Chronic nonalcoholic liver disease | Gastrointestinal | 10,142 / 329,353 | 0.96 (0.94–0.98) | -0.045 (0.010) | 8.70e-06 |

Sensitivity analysis in which the standard whole-cohort logistic model (ordinal APOE genotype: ε2/ε2 < ε2/ε3 < ε3/ε3 < ε2/ε4 < ε3/ε4 < ε4/ε4; adjusted for age, sex, 16 genetic PCs and EHR length) was additionally adjusted for social determinants of health. Results are shown for the 27 phecodes that reached Bonferroni-corrected significance in the primary whole-cohort PheWAS, ordered by ascending P value in that primary analysis. All 27 phecodes retained the same direction of effect and remained highly significant, indicating robustness of the primary findings to SDOH adjustment. OR, odds ratio; CI, confidence interval; β, log-odds effect size; SE, standard error.

**Table S3.** Phenome-wide association results for APOE ε2 allele dosage.

| Phecode | Phenotype | Category | Cases / Controls | OR (95% CI) | β (SE) | P |
|---|---|---|---|---|---|---|
| EM_239 | Hyperlipidemia | Endocrine/Metab | 143,368 / 196,251 | 0.71 (0.69–0.72) | -0.348 (0.011) | 1.69e-205 |
| EM_239.1 | Hypercholesterolemia | Endocrine/Metab | 79,851 / 265,403 | 0.74 (0.72–0.76) | -0.303 (0.013) | 3.07e-128 |
| EM_239.11 | Pure hypercholesterolemia | Endocrine/Metab | 45,995 / 302,961 | 0.72 (0.70–0.74) | -0.327 (0.016) | 1.52e-97 |
| EM_239.21 | Pure hyperglyceridemia | Endocrine/Metab | 8,322 / 354,304 | 1.53 (1.45–1.61) | 0.427 (0.027) | 9.44e-58 |
| EM_239.3 | Mixed hyperlipidemia | Endocrine/Metab | 45,183 / 308,966 | 0.82 (0.80–0.85) | -0.196 (0.015) | 8.51e-40 |
| CV_436 | Atherosclerosis [ASCVD] | Cardiovascular | 45,807 / 305,469 | 0.88 (0.86–0.91) | -0.123 (0.015) | 1.88e-16 |
| CV_404.2 | Coronary atherosclerosis [Atherosclerotic heart disease] | Cardiovascular | 39,668 / 315,052 | 0.89 (0.86–0.91) | -0.120 (0.016) | 2.19e-14 |
| CV_404 | Ischemic heart disease | Cardiovascular | 44,858 / 308,158 | 0.90 (0.88–0.93) | -0.103 (0.015) | 2.90e-12 |

| Phecode | Phenotype | Category | Cases / Controls | OR (95% CI) | β (SE) | P |
|---|---|---|---|---|---|---|
| EM_239.2 | Hyperglyceridemia | Endocrine/Metab | 50,899 / 301,143 | 0.91 (0.89–0.94) | -0.093 (0.014) | 1.29e-11 |
| CV_433.2 | Occlusion and stenosis of precerebral arteries | Cardiovascular | 8,198 / 352,946 | 0.86 (0.80–0.91) | -0.155 (0.032) | 9.75e-07 |
| EM_244 | Disorders of lipoprotein metabolism and other lipidemias | Endocrine/Metab | 4,184 / 358,110 | 0.80 (0.73–0.88) | -0.223 (0.046) | 1.02e-06 |
| CV_413.13 | Mitral valve stenosis | Cardiovascular | 712 / 366,437 | 1.49 (1.26–1.76) | 0.398 (0.085) | 2.44e-06 |
| NS_328.11 | Alzheimer's disease | Neurological | 1,030 / 366,205 | 0.62 (0.50–0.76) | -0.486 (0.110) | 1.05e-05 |

Phenome-wide association results for APOE ε2 allele dosage in the whole cohort, estimated from a joint model that adjusts for ε4 allele dosage (in addition to the standard covariates). Of 2,627 tested phecodes, 13 surpassed the Bonferroni-corrected significance threshold ($P < 1.90\times10^{-5}$). Results are sorted by ascending P value. OR, odds ratio per additional ε2 allele conditional on ε4 dosage; CI, confidence interval; β, log-odds effect size; SE, standard error.

**Table S4.** Phenome-wide association results for APOE ε4 allele dosage.

| Phecode | Phenotype | Category | Cases / Controls | OR (95% CI) | β (SE) | P |
|---|---|---|---|---|---|---|
| EM_239 | Hyperlipidemia | Endocrine/Metab | 143,368 / 196,251 | 1.20 (1.18–1.22) | 0.182 (0.008) | $1.01\times10^{-102}$ |
| NS_328.1 | Dementias | Neurological | 4,021 / 361,476 | 1.72 (1.63–1.82) | 0.544 (0.029) | $1.18\times10^{-78}$ |
| NS_328.11 | Alzheimer's disease | Neurological | 1,030 / 366,205 | 2.60 (2.35–2.89) | 0.957 (0.052) | $3.93\times10^{-75}$ |
| NS_328 | Dementias and cerebral degeneration | Neurological | 4,859 / 358,742 | 1.62 (1.53–1.70) | 0.480 (0.027) | $8.69\times10^{-72}$ |
| NS_329.1 | Memory loss | Neurological | 13,036 / 346,620 | 1.32 (1.28–1.36) | 0.277 (0.018) | $2.05\times10^{-56}$ |
| EM_239.1 | Hypercholesterolemia | Endocrine/Metab | 79,851 / 265,403 | 1.15 (1.13–1.17) | 0.142 (0.009) | $2.53\times10^{-55}$ |
| EM_239.11 | Pure hypercholesterolemia | Endocrine/Metab | 45,995 / 302,961 | 1.16 (1.14–1.19) | 0.149 (0.011) | $6.28\times10^{-43}$ |
| NS_329.5 | Mild cognitive impairment | Neurological | 5,005 / 360,091 | 1.45 (1.37–1.53) | 0.370 (0.027) | $4.08\times10^{-42}$ |
| NS_329 | Symptoms and signs involving cognitive functions and awareness | Neurological | 28,556 / 322,629 | 1.18 (1.15–1.21) | 0.167 (0.013) | $7.96\times10^{-41}$ |
| EM_239.2 | Hyperglyceridemia | Endocrine/Metab | 50,899 / 301,143 | 1.12 (1.10–1.15) | 0.117 (0.010) | $6.76\times10^{-31}$ |
| EM_239.3 | Mixed hyperlipidemia | Endocrine/Metab | 45,183 / 308,966 | 1.11 (1.09–1.13) | 0.105 (0.011) | $2.70\times10^{-23}$ |

| | | | | | | |
|---|---|---|---|---|---|---|
| GI_542.2 | Fatty liver disease (FLD) | Gastrointestinal | 18,234 / 337,190 | 0.86 (0.84–0.89) | -0.147 (0.017) | $1.17 \times 10^{-18}$ |
| GI_542 | Chronic liver disease | Gastrointestinal | 26,591 / 326,039 | 0.90 (0.87–0.92) | -0.111 (0.014) | $7.95 \times 10^{-16}$ |
| EM_239.21 | Pure hyperglyceridemia | Endocrine/Metab | 8,322 / 354,304 | 1.15 (1.10–1.21) | 0.142 (0.023) | $4.10 \times 10^{-10}$ |
| NS_328.12 | Vascular dementia | Neurological | 591 / 366,794 | 1.57 (1.36–1.82) | 0.451 (0.075) | $1.87 \times 10^{-9}$ |
| EM_249.1 | Cerebral amyloid angiopathy* | Endocrine/Metab | 101 / 367,589 | 2.59 (1.87–3.60) | 0.953 (0.167) | $1.19 \times 10^{-8}$ |
| NS_329.8 | Altered mental status, unspecified | Neurological | 7,045 / 351,898 | 1.13 (1.08–1.19) | 0.125 (0.024) | $1.48 \times 10^{-7}$ |
| GI_542.7 | Chronic nonalcoholic liver disease | Gastrointestinal | 11,027 / 349,718 | 0.90 (0.86–0.93) | -0.109 (0.021) | $2.00 \times 10^{-7}$ |
| MB_308.4 | Neurocognitive disorder* | Mental | 167 / 367,393 | 1.96 (1.52–2.54) | 0.674 (0.131) | $2.70 \times 10^{-7}$ |
| CV_404.2 | Coronary atherosclerosis [Atherosclerotic heart disease] | Cardiovascular | 39,668 / 315,052 | 1.06 (1.04–1.09) | 0.060 (0.012) | $4.21 \times 10^{-7}$ |
| NS_328.14 | Dementia with Lewy bodies | Neurological | 138 / 367,537 | 2.08 (1.56–2.78) | 0.733 (0.148) | $7.37 \times 10^{-7}$ |
| CV_404 | Ischemic heart disease | Cardiovascular | 44,858 / 308,158 | 1.05 (1.03–1.08) | 0.052 (0.011) | $2.86 \times 10^{-6}$ |

Phenome-wide association results for APOE ε4 allele dosage in the whole cohort, estimated from a joint model that adjusts for ε2 allele dosage (in addition to the standard covariates). Of 2,627 tested phecodes, 22 surpassed the Bonferroni-corrected significance threshold ($P < 1.90 \times 10^{-}5$). Results are sorted by ascending P value. OR, odds ratio per additional ε4 allele conditional on ε2 dosage; CI, confidence interval; β, log-odds effect size; SE, standard error.

### **Table S5.** Diplotype-level odds ratios for 26 significant whole-cohort PheWAS phecodes.

| Phecode | Phenotype | Category | N cases | ε2/ε2 | ε2/ε3 | ε2/ε4 | ε3/ε4 | ε4/ε4 |
|---|---|---|---|---|---|---|---|---|
| EM_239 | Hyperlipidemia | Endocrine/Metab | 143,368 | 0.75 (0.68–0.82) | 0.71 (0.69–0.73) | 0.85 (0.81–0.90) | 1.18 (1.16–1.21) | 1.35 (1.28–1.42) |
| EM_239.1 | Hypercholesterolemia | Endocrine/Metab | 79,851 | 0.76 (0.68–0.85) | 0.73 (0.71–0.76) | 0.86 (0.81–0.92) | 1.14 (1.12–1.17) | 1.26 (1.19–1.33) |
| EM_239.11 | Pure hypercholesterolemia | Endocrine/Metab | 45,995 | 0.67 (0.58–0.77) | 0.72 (0.69–0.75) | 0.85 (0.79–0.91) | 1.15 (1.12–1.18) | 1.29 (1.20–1.38) |

| Phecode | Phenotype | Category | N cases | ε2/ε2 | ε2/ε3 | ε2/ε4 | ε3/ε4 | ε4/ε4 |
|---|---|---|---|---|---|---|---|---|
| NS_328.1 | Dementias | Neurological | 4,021 | 0.85 (0.54–1.33) | 0.86 (0.77–0.96) | 1.14 (0.92–1.41) | 1.65 (1.53–1.78) | 3.45 (2.96–4.01) |
| NS_328.11 | Alzheimer's disease | Neurological | 1,030 | 0.60 (0.19–1.88) | 0.70 (0.54–0.90) | 0.93 (0.56–1.56) | 2.52 (2.19–2.89) | 7.73 (6.07–9.86) |
| NS_328 | Dementias and cerebral degeneration | Neurological | 4,859 | 0.83 (0.55–1.24) | 0.90 (0.81–0.99) | 1.15 (0.94–1.39) | 1.57 (1.47–1.68) | 2.94 (2.54–3.40) |
| NS_329.1 | Memory loss | Neurological | 13,036 | 1.01 (0.81–1.27) | 0.92 (0.86–0.97) | 1.07 (0.95–1.21) | 1.30 (1.24–1.35) | 1.87 (1.68–2.08) |
| EM_239.3 | Mixed hyperlipidemia | Endocrine/Metab | 45,183 | 0.93 (0.82–1.06) | 0.80 (0.77–0.83) | 0.92 (0.86–0.99) | 1.11 (1.08–1.14) | 1.18 (1.10–1.27) |
| EM_239.2 | Hyperglyceridemia | Endocrine/Metab | 50,899 | 1.13 (1.01–1.28) | 0.89 (0.86–0.91) | 1.01 (0.94–1.08) | 1.12 (1.10–1.15) | 1.22 (1.14–1.30) |
| NS_329 | Symptoms and signs involving cognitive functions and awareness | Neurological | 28,556 | 0.92 (0.79–1.09) | 0.96 (0.92–1.00) | 1.03 (0.94–1.12) | 1.16 (1.13–1.20) | 1.52 (1.41–1.65) |
| NS_329.5 | Mild cognitive impairment | Neurological | 5,005 | 1.19 (0.85–1.66) | 0.90 (0.82–0.99) | 1.24 (1.03–1.49) | 1.41 (1.32–1.51) | 2.26 (1.93–2.65) |
| GI_542.2 | Fatty liver disease (FLD) | Gastrointestinal | 18,234 | 0.94 (0.78–1.15) | 0.99 (0.95–1.04) | 0.88 (0.78–0.98) | 0.85 (0.82–0.88) | 0.82 (0.73–0.92) |
| CV_404.2 | Coronary atherosclerosis [Atherosclerotic heart disease] | Cardiovascular | 39,668 | 0.88 (0.77–1.02) | 0.88 (0.85–0.91) | 0.95 (0.88–1.03) | 1.05 (1.02–1.08) | 1.16 (1.07–1.25) |
| GI_542 | Chronic liver disease | Gastrointestinal | 26,591 | 0.96 (0.81–1.13) | 1.00 (0.96–1.04) | 0.91 (0.84–1.00) | 0.88 (0.86–0.91) | 0.86 (0.78–0.94) |
| CV_436 | Atherosclerosis [ASCVD] | Cardiovascular | 45,807 | 0.89 (0.78–1.02) | 0.88 (0.85–0.91) | 0.93 (0.86–1.00) | 1.03 (1.01–1.06) | 1.14 (1.05–1.22) |
| CV_404 | Ischemic heart disease | Cardiovascular | 44,858 | 0.89 (0.78–1.02) | 0.90 (0.87–0.93) | 0.95 (0.88–1.02) | 1.04 (1.02–1.07) | 1.16 (1.08–1.25) |
| NS_328.12 | Vascular dementia | Neurological | 591 | 0.55 (0.14–2.20) | 0.92 (0.70–1.21) | 0.97 (0.54–1.73) | 1.55 (1.29–1.88) | 2.80 (1.88–4.18) |
| NS_329.8 | Altered mental status, unspecified | Neurological | 7,045 | 0.92 (0.68–1.25) | 0.97 (0.90–1.05) | 0.88 (0.74–1.04) | 1.13 (1.06–1.19) | 1.42 (1.23–1.64) |
| CV_403 | Angina pectoris | Cardiovascular | 14,293 | 1.03 (0.84–1.27) | 0.90 (0.85–0.95) | 0.97 (0.86–1.09) | 1.07 (1.02–1.11) | 1.20 (1.06–1.34) |

| Phecode | Phenotype | Category | N cases | ε2/ε2 | ε2/ε3 | ε2/ε4 | ε3/ε4 | ε4/ε4 |
|---|---|---|---|---|---|---|---|---|
| CV_414.5 | Ischemic cardiomyopathy* | Cardiovascular | 3,080 | 0.59 (0.33–1.05) | 0.82 (0.72–0.93) | 1.01 (0.79–1.29) | 1.17 (1.08–1.28) | 1.12 (0.88–1.44) |
| EM_244 | Disorders of lipoprotein metabolism and other lipidemias | Endocrine/Metab | 4,184 | 1.01 (0.70–1.47) | 0.78 (0.70–0.86) | 0.79 (0.63–1.01) | 1.12 (1.04–1.21) | 1.19 (0.96–1.48) |
| CV_433.2 | Occlusion and stenosis of precerebral arteries | Cardiovascular | 8,198 | 0.78 (0.58–1.05) | 0.84 (0.78–0.90) | 0.98 (0.84–1.14) | 1.05 (1.00–1.11) | 1.12 (0.95–1.31) |
| CV_433 | Other cerebrovascular disease | Cardiovascular | 14,963 | 0.95 (0.78–1.17) | 0.88 (0.83–0.93) | 0.99 (0.89–1.11) | 1.04 (0.99–1.08) | 1.13 (1.01–1.27) |
| NS_328.14 | Dementia with Lewy bodies | Neurological | 138 | 1.55 (0.21–11.19) | 1.07 (0.60–1.91) | 1.89 (0.68–5.19) | 2.24 (1.54–3.27) | 3.85 (1.66–8.94) |
| GE_965.4 | Familial hypercholesterolemia* | Genetic | 947 | 1.29 (0.64–2.60) | 0.66 (0.52–0.84) | 0.96 (0.61–1.50) | 1.27 (1.09–1.48) | 1.07 (0.68–1.70) |
| GI_542.7 | Chronic nonalcoholic liver disease | Gastrointestinal | 11,027 | 0.89 (0.69–1.15) | 0.96 (0.90–1.02) | 0.93 (0.81–1.07) | 0.88 (0.84–0.93) | 0.83 (0.72–0.96) |

Each cell reports the odds ratio (95% confidence interval) for the indicated APOE diplotype relative to the ε3/ε3 reference, for the 26 phecodes reaching Bonferroni-corrected significance in the whole-cohort PheWAS for which stable estimates could be obtained across all five non-reference diplotypes. N cases is the number of cases contributing to each phecode (constant across diplotype comparisons). MB_308.4 (Neurocognitive disorder), which reached significance in the additive whole-cohort analysis, is not shown because at least one diplotype cell (ε2/ε2) had insufficient counts for a stable estimate, so it could not be tested.

**Table S6.** APOE genotype associations reaching Bonferroni significance in female participants.

| Phecode | Phenotype | Category | Cases / Controls | OR (95% CI) | β (SE) | P |
|---|---|---|---|---|---|---|
| EM_239 | Hyperlipidemia | Endocrine/Metab | 79,036 / 128,944 | 1.17 (1.16–1.18) | 0.159 (0.005) | 8.16e-208 |
| EM_239.1 | Hypercholesterolemia | Endocrine/Metab | 44,825 / 167,039 | 1.14 (1.13–1.16) | 0.135 (0.006) | 2.40e-127 |
| EM_239.11 | Pure hypercholesterolemia | Endocrine/Metab | 26,495 / 187,621 | 1.15 (1.14–1.17) | 0.141 (0.007) | 4.64e-97 |
| EM_239.3 | Mixed hyperlipidemia | Endocrine/Metab | 24,558 / 192,720 | 1.10 (1.09–1.12) | 0.097 (0.007) | 9.19e-48 |

| Phecode | Phenotype | Category | Cases / Controls | OR (95% CI) | β (SE) | P |
|---|---|---|---|---|---|---|
| NS_328.1 | Dementias | Neurological | 2,046 / 221,643 | 1.32 (1.27–1.38) | 0.279 (0.021) | 4.65e-42 |
| NS_328.11 | Alzheimer's disease | Neurological | 550 / 224,077 | 1.68 (1.56–1.81) | 0.520 (0.039) | 1.43e-41 |
| NS_328 | Dementias and cerebral degeneration | Neurological | 2,430 / 220,331 | 1.28 (1.23–1.33) | 0.245 (0.019) | 2.34e-38 |
| EM_239.2 | Hyperglyceridemia | Endocrine/Metab | 27,534 / 188,613 | 1.08 (1.07–1.10) | 0.082 (0.006) | 5.64e-37 |
| NS_329.1 | Memory loss | Neurological | 8,180 / 211,703 | 1.14 (1.12–1.17) | 0.135 (0.011) | 4.27e-36 |
| NS_329.5 | Mild cognitive impairment | Neurological | 2,337 / 221,291 | 1.22 (1.18–1.27) | 0.201 (0.019) | 2.78e-25 |
| NS_329 | Symptoms and signs involving cognitive functions and awareness | Neurological | 16,417 / 198,841 | 1.08 (1.07–1.10) | 0.082 (0.008) | 2.98e-25 |
| CV_436 | Atherosclerosis [ASCVD] | Cardiovascular | 19,526 / 196,516 | 1.05 (1.03–1.06) | 0.045 (0.008) | 4.84e-09 |
| CV_404.2 | Coronary atherosclerosis [Atherosclerotic heart disease] | Cardiovascular | 15,817 / 202,240 | 1.05 (1.03–1.07) | 0.048 (0.008) | 8.07e-09 |
| GI_542.2 | Fatty liver disease (FLD) | Gastrointestinal | 11,055 / 206,468 | 0.95 (0.93–0.97) | -0.054 (0.010) | 3.63e-08 |
| GI_542 | Chronic liver disease | Gastrointestinal | 15,302 / 200,706 | 0.96 (0.94–0.97) | -0.046 (0.008) | 4.81e-08 |
| EM_244 | Disorders of lipoprotein metabolism and other lipidemias | Endocrine/Metab | 2,305 / 219,549 | 1.10 (1.06–1.15) | 0.099 (0.020) | 7.71e-07 |
| CV_404 | Ischemic heart disease | Cardiovascular | 18,599 / 198,356 | 1.04 (1.02–1.05) | 0.037 (0.008) | 1.64e-06 |

| Phecode | Phenotype | Category | Cases / Controls | OR (95% CI) | β (SE) | P |
|---|---|---|---|---|---|---|
| NS_329.8 | Altered mental status, unspecified | Neurological | 3,641 / 216,644 | 1.08 (1.04–1.11) | 0.073 (0.016) | 3.19e-06 |
| GE_965.4 | Familial hypercholesterolemia* | Genetic | 597 / 223,992 | 1.19 (1.11–1.28) | 0.174 (0.038) | 3.93e-06 |
| CV_433.2 | Occlusion and stenosis of precerebral arteries | Cardiovascular | 4,092 / 217,445 | 1.07 (1.04–1.11) | 0.070 (0.015) | 4.91e-06 |
| NS_346.3 | Anoxic brain damage | Neurological | 128 / 224,646 | 1.42 (1.22–1.65) | 0.349 (0.078) | 7.59e-06 |
| SO_374.511 | Nonexuadative (dry) age-related macular degeneration | Sense organs | 1,791 / 222,401 | 0.90 (0.85–0.94) | -0.108 (0.025) | 2.02e-05 |

Of 2,384 tested phecodes, 22 surpassed the stratum-specific Bonferroni-corrected significance threshold ($P < 2.10\times10^{-5}$) in female participants (sex assigned at birth). The ordinal APOE genotype (ε2/ε2 < ε2/ε3 < ε3/ε3 < ε2/ε4 < ε3/ε4 < ε4/ε4) was modelled with logistic regression adjusted for age, 16 genetic PCs and EHR length. Results are sorted by ascending P value. OR, odds ratio; CI, confidence interval; β, log-odds effect size; SE, standard error.

**Table S7.** APOE genotype associations reaching Bonferroni significance in male participants.

| Phecode | Phenotype | Category | Cases / Controls | OR (95% CI) | β (SE) | P |
|---|---|---|---|---|---|---|
| EM_239 | Hyperlipidemia | Endocrine/Metab | 64,332 / 67,307 | 1.12 (1.11–1.13) | 0.112 (0.006) | 3.36e-68 |
| NS_328.1 | Dementias | Neurological | 1,975 / 139,833 | 1.31 (1.26–1.36) | 0.269 (0.021) | 1.22e-37 |
| EM_239.1 | Hypercholesterolemia | Endocrine/Metab | 35,026 / 98,364 | 1.09 (1.07–1.10) | 0.083 (0.007) | 1.74e-35 |
| NS_328.11 | Alzheimer's disease | Neurological | 480 / 142,128 | 1.66 (1.53–1.80) | 0.508 (0.041) | 4.17e-35 |
| NS_328 | Dementias and cerebral degeneration | Neurological | 2,429 / 138,411 | 1.26 (1.22–1.31) | 0.233 (0.019) | 1.82e-34 |
| EM_239.11 | Pure hypercholesterolemia | Endocrine/Metab | 19,500 / 115,340 | 1.09 (1.07–1.11) | 0.087 (0.008) | 1.43e-27 |
| NS_329.1 | Memory loss | Neurological | 4,856 / 134,917 | 1.15 (1.12–1.19) | 0.142 (0.014) | 2.79e-24 |
| NS_329 | Symptoms and signs involving cognitive functions and awareness | Neurological | 12,139 / 123,788 | 1.08 (1.07–1.10) | 0.081 (0.009) | 3.72e-18 |

| Phecode | Phenotype | Category | Cases / Controls | OR (95% CI) | β (SE) | P |
|---|---|---|---|---|---|---|
| NS_329.5 | Mild cognitive impairment | Neurological | 2,668 / 138,800 | 1.17 (1.13–1.21) | 0.158 (0.019) | 2.80e-17 |
| EM_239.3 | Mixed hyperlipidemia | Endocrine/Metab | 20,625 / 116,246 | 1.06 (1.04–1.07) | 0.056 (0.008) | 1.86e-13 |
| EM_239.2 | Hyperglyceridemia | Endocrine/Metab | 23,365 / 112,530 | 1.05 (1.04–1.07) | 0.051 (0.007) | 3.91e-12 |
| GI_542.2 | Fatty liver disease (FLD) | Gastrointestinal | 7,179 / 130,722 | 0.92 (0.90–0.95) | -0.080 (0.012) | 1.07e-10 |
| CV_404.2 | Coronary atherosclerosis [Atherosclerotic heart disease] | Cardiovascular | 23,851 / 112,812 | 1.05 (1.03–1.06) | 0.045 (0.007) | 1.32e-09 |
| CV_404 | Ischemic heart disease | Cardiovascular | 26,259 / 109,802 | 1.04 (1.03–1.06) | 0.043 (0.007) | 2.57e-09 |
| GI_542 | Chronic liver disease | Gastrointestinal | 11,289 / 125,333 | 0.95 (0.93–0.97) | -0.053 (0.010) | 6.58e-08 |
| CV_436 | Atherosclerosis [ASCVD] | Cardiovascular | 26,281 / 108,953 | 1.04 (1.02–1.05) | 0.036 (0.007) | 6.81e-07 |
| NS_328.12 | Vascular dementia | Neurological | 319 / 142,312 | 1.28 (1.16–1.41) | 0.245 (0.051) | 1.66e-06 |
| CV_419.2 | Presence of cardiac defibrillator | Cardiovascular | 1,822 / 140,509 | 1.11 (1.06–1.15) | 0.101 (0.022) | 5.04e-06 |
| NS_336.4 | Mononeuritis of upper limb | Neurological | 8,708 / 130,252 | 0.95 (0.93–0.97) | -0.051 (0.011) | 5.46e-06 |

Of 2,066 tested phecodes, 19 surpassed the stratum-specific Bonferroni-corrected significance threshold ($P < 2.42\times10^{-5}$) in male participants (sex assigned at birth). The ordinal APOE genotype (ε2/ε2 < ε2/ε3 < ε3/ε3 < ε2/ε4 < ε3/ε4 < ε4/ε4) was modelled with logistic regression adjusted for age, 16 genetic PCs and EHR length. Results are sorted by ascending P value. OR, odds ratio; CI, confidence interval; β, log-odds effect size; SE, standard error.

**Table S8.** APOE × sex interaction results for the 5 phecodes with significant sex heterogeneity.

| Phecode | Phenotype | Category | Cases / Controls | β Female | β Male | β Interaction (SE) | P (interaction) | P (FDR) |
|---|---|---|---|---|---|---|---|---|
| EM_239 | Hyperlipidemia | Endocrine/Metab | 143,368 / 196,251 | +0.165 | +0.102 | -0.063 (0.008) | 8.67e-15 | 4.34e-14 |
| EM_239.1 | Hypercholesterolemia | Endocrine/Metab | 79,851 / 265,403 | +0.136 | +0.079 | -0.058 (0.009) | 2.46e-11 | 6.15e-11 |
| EM_239.11 | Pure hypercholesterolemia | Endocrine/Metab | 45,995 / 302,961 | +0.141 | +0.086 | -0.055 (0.010) | 1.10e-07 | 1.83e-07 |
| EM_239.3 | Mixed hyperlipidemia | Endocrine/Metab | 45,183 / 308,966 | +0.099 | +0.053 | -0.047 (0.010) | 3.67e-06 | 4.59e-06 |

| Phecode | Phenotype | Category | Cases / Controls | β Female | β Male | β Interaction (SE) | P (interaction) | P (FDR) |
|---|---|---|---|---|---|---|---|---|
| EM_239.2 | Hyperglyceridemia | Endocrine/Metab | 50,899 / 301,143 | +0.083 | +0.047 | -0.036 (0.010) | 1.75e-04 | 1.75e-04 |

Sex-specific log-odds effect sizes (β Female, β Male) are shown alongside the APOE × sex interaction coefficient (β Interaction) with its standard error, the raw interaction P value, and the Benjamini–Hochberg FDR-adjusted P value. The interaction term is parameterised such that a negative coefficient indicates a smaller APOE effect in males relative to females. All 5 FDR-significant interactions fall within the hyperlipidemia phecode hierarchy and coincide with the phecodes that were also Bonferroni-significant under Cochran's Q heterogeneity testing.

**Table S9**. Ancestry-stratified PheWAS results for APOE across four ancestral groups.

| Ancestry | Phecode | Phenotype | Category | Cases / Controls | OR (95% CI) | β (SE) | P |
|---|---|---|---|---|---|---|---|
| **AFR** | EM_239 | Hyperlipidemia | Endocrine/Metab | 20,900 / 38,978 | 1.12 (1.11–1.14) | +0.116 (0.008) | 4.92e-44 |
| | EM_239.1 | Hypercholesterolemia | Endocrine/Metab | 10,382 / 50,493 | 1.11 (1.09–1.13) | +0.104 (0.010) | 3.64e-26 |
| | EM_239.11 | Pure hypercholesterolemia | Endocrine/Metab | 6,075 / 55,381 | 1.12 (1.10–1.15) | +0.115 (0.012) | 1.20e-21 |
| | EM_239.3 | Mixed hyperlipidemia | Endocrine/Metab | 5,707 / 56,481 | 1.09 (1.06–1.11) | +0.083 (0.012) | 4.73e-12 |
| | EM_239.2 | Hyperglyceridemia | Endocrine/Metab | 6,188 / 55,723 | 1.08 (1.06–1.11) | +0.078 (0.012) | 2.06e-11 |
| | NS_328.1 | Dementias | Neurological | 609 / 63,266 | 1.24 (1.16–1.33) | +0.217 (0.034) | 2.33e-10 |
| | NS_328 | Dementias and cerebral degeneration | Neurological | 755 / 62,818 | 1.20 (1.13–1.27) | +0.179 (0.031) | 4.95e-09 |
| | NS_328.11 | Alzheimer's disease | Neurological | 113 / 64,081 | 1.45 (1.24–1.70) | +0.374 (0.080) | 3.16e-06 |
| | NS_329.1 | Memory loss | Neurological | 1,679 / 61,387 | 1.10 (1.06–1.15) | +0.096 (0.021) | 4.35e-06 |
| | CV_414.5 | Ischemic cardiomyopathy* | Cardiovascular | 497 / 63,443 | 1.17 (1.09–1.26) | +0.158 (0.037) | 2.13e-05 |
| **AMR** | EM_239 | Hyperlipidemia | Endocrine/Metab | 20,759 / 41,773 | 1.11 (1.09–1.14) | +0.107 (0.011) | 5.87e-23 |
| | EM_239.1 | Hypercholesterolemia | Endocrine/Metab | 11,066 / 52,641 | 1.08 (1.05–1.10) | +0.073 (0.012) | 3.11e-09 |
| | EM_239.11 | Pure hypercholesterolemia | Endocrine/Metab | 5,979 / 58,184 | 1.08 (1.05–1.12) | +0.081 (0.015) | 1.49e-07 |
| | MB_280.1 | Alcohol use disorders | Mental | 4,062 / 61,005 | 1.09 (1.05–1.12) | +0.082 (0.017) | 2.01e-06 |

| Ancestry | Phecode | Phenotype | Category | Cases / Controls | OR (95% CI) | β (SE) | P |
|---|---|---|---|---|---|---|---|
| | NS_328.11 | Alzheimer's disease | Neurological | 141 / 66,625 | 1.45 (1.23–1.71) | +0.374 (0.083) | 7.44e-06 |
| | GI_542.2 | Fatty liver disease (FLD) | Gastrointestinal | 4,873 / 58,709 | 0.93 (0.90–0.96) | -0.074 (0.017) | 1.06e-05 |
| **EAS** | EM_239 | Hyperlipidemia | Endocrine/Metab | 2,879 / 6,031 | 1.20 (1.13–1.27) | +0.183 (0.029) | 5.40e-10 |
| | EM_239.1 | Hypercholesterolemia | Endocrine/Metab | 1,630 / 7,516 | 1.15 (1.08–1.23) | +0.144 (0.032) | 7.16e-06 |
| **EUR** | EM_239 | Hyperlipidemia | Endocrine/Metab | 97,190 / 106,271 | 1.17 (1.16–1.18) | +0.158 (0.005) | 1.88e-201 |
| | EM_239.1 | Hypercholesterolemia | Endocrine/Metab | 55,789 / 150,805 | 1.13 (1.12–1.14) | +0.124 (0.005) | 6.70e-121 |
| | EM_239.11 | Pure hypercholesterolemia | Endocrine/Metab | 32,528 / 176,653 | 1.14 (1.12–1.15) | +0.127 (0.006) | 2.01e-91 |
| | NS_328.1 | Dementias | Neurological | 2,641 / 217,903 | 1.39 (1.34–1.44) | +0.326 (0.018) | 6.38e-73 |
| | NS_328.11 | Alzheimer's disease | Neurological | 741 / 220,809 | 1.77 (1.66–1.89) | +0.571 (0.033) | 2.09e-67 |
| | NS_328 | Dementias and cerebral degeneration | Neurological | 3,212 / 216,029 | 1.33 (1.29–1.37) | +0.284 (0.017) | 2.52e-66 |
| | NS_329.1 | Memory loss | Neurological | 9,108 / 207,200 | 1.18 (1.16–1.21) | +0.167 (0.010) | 5.88e-60 |
| | NS_329.5 | Mild cognitive impairment | Neurological | 3,701 / 216,356 | 1.25 (1.21–1.29) | +0.221 (0.016) | 1.15e-45 |
| | EM_239.3 | Mixed hyperlipidemia | Endocrine/Metab | 31,401 / 181,105 | 1.09 (1.07–1.10) | +0.084 (0.006) | 1.60e-42 |
| | NS_329 | Symptoms and signs involving cognitive functions and awareness | Neurological | 19,734 / 191,535 | 1.10 (1.09–1.12) | +0.100 (0.007) | 1.82e-42 |
| | EM_239.2 | Hyperglyceridemia | Endocrine/Metab | 35,039 / 176,185 | 1.07 (1.06–1.09) | +0.071 (0.006) | 1.88e-33 |
| | GI_542.2 | Fatty liver disease (FLD) | Gastrointestinal | 10,667 / 204,173 | 0.93 (0.91–0.95) | -0.073 (0.010) | 2.87e-13 |
| | GI_542 | Chronic liver disease | Gastrointestinal | 15,294 / 197,814 | 0.94 (0.93–0.96) | -0.060 (0.008) | 7.42e-13 |
| | CV_404.2 | Coronary atherosclerosis [Atherosclerotic heart disease] | Cardiovascular | 28,166 / 185,053 | 1.05 (1.03–1.06) | +0.047 (0.007) | 3.18e-12 |

| Ancestry | Phecode | Phenotype | Category | Cases / Controls | OR (95% CI) | β (SE) | P |
|---|---|---|---|---|---|---|---|
| | CV_436 | Atherosclerosis [ASCVD] | Cardiovascular | 32,251 / 178,777 | 1.04 (1.03–1.06) | +0.043 (0.007) | 3.42e-11 |
| | CV_404 | Ischemic heart disease | Cardiovascular | 30,905 / 181,510 | 1.04 (1.03–1.06) | +0.043 (0.007) | 5.79e-11 |
| | NS_328.12 | Vascular dementia | Neurological | 356 / 221,330 | 1.30 (1.18–1.43) | +0.263 (0.049) | 7.03e-08 |
| | EM_244 | Disorders of lipoprotein metabolism and other lipidemias | Endocrine/Metab | 3,315 / 214,434 | 1.09 (1.06–1.13) | +0.090 (0.017) | 1.97e-07 |
| | NS_329.8 | Altered mental status, unspecified | Neurological | 4,543 / 212,110 | 1.07 (1.04–1.11) | +0.072 (0.014) | 7.22e-07 |
| | CV_433 | Other cerebrovascular disease | Cardiovascular | 10,138 / 204,814 | 1.05 (1.03–1.07) | +0.051 (0.010) | 7.65e-07 |
| | MB_308.4 | Neurocognitive disorder* | Mental | 117 / 221,642 | 1.48 (1.26–1.74) | +0.394 (0.082) | 1.35e-06 |
| | CV_403 | Angina pectoris | Cardiovascular | 9,769 / 206,934 | 1.05 (1.03–1.07) | +0.048 (0.010) | 4.44e-06 |
| | NS_328.14 | Dementia with Lewy bodies | Neurological | 107 / 221,735 | 1.48 (1.25–1.76) | +0.394 (0.087) | 5.34e-06 |

Bonferroni-significant associations from ancestry-stratified PheWAS in European (EUR; 2,272 phecodes tested, $\alpha = 2.20 \times 10^{-5}$), African (AFR; 1,824 tested, $\alpha = 2.74 \times 10^{-5}$), Admixed American (AMR; 1,843 tested, $\alpha = 2.71 \times 10^{-5}$), and East Asian (EAS; 901 tested, $\alpha = 5.55 \times 10^{-5}$) participants. In each stratum, the ordinal APOE genotype (ε2/ε2 < ε2/ε3 < ε3/ε3 < ε2/ε4 < ε3/ε4 < ε4/ε4) was modelled with logistic regression adjusted for age, sex at birth, 16 genetic PCs, and EHR length. South Asian (SAS; 486 tested) and Middle Eastern (MID; 335 tested) strata yielded no Bonferroni-significant associations and are not shown. Rows are grouped by ancestry, then sorted by ascending raw P value within ancestry.

### **Table S10.** Ancestry heterogeneity sensitivity analysis.

| | | Per-ancestry effect — OR (case N) | | | | Heterogeneity: 3 ancestries (EUR/AFR/AMR) | | | Heterogeneity: 4 ancestries (EUR/AFR/AMR/EAS) | | | |
|---|---|---|---|---|---|---|---|---|---|---|---|---|
| **Phecode** | **Description** | **EUR** | **AFR** | **AMR** | **EAS** | ***Q*** | ***p*-FDR** | ***I*² (%)** | ***Q*** | ***p*-FDR** | ***I*² (%)** | **Status** |
| EM_239 | Hyperlipidemia | 1.171 (97,190) | 1.123 (20,900) | 1.112 (20,759) | 1.200 (2,879) | 28.02 | **$1.28\times10^{-5}$** | 92.9 | 30.07 | **$3.07\times10^{-5}$** | 90.0 | consistent_HET |
| NS_328.1 | Dementias | 1.386 (2,641) | 1.242 (609) | 1.115 (665) | 1.195 (64) | 27.16 | **$1.28\times10^{-5}$** | 92.6 | 27.75 | **$4.72\times10^{-5}$** | 89.2 | consistent_HET |
| NS_329.1 | Memory loss | 1.182 (9,108) | 1.101 (1,679) | 1.043 (1,872) | 1.146 (244) | 26.49 | **$1.28\times10^{-5}$** | 92.5 | 26.49 | **$4.92\times10^{-5}$** | 88.7 | consistent_HET |
| NS_329.5 | Mild cognitive impairment | 1.248 (3,701) | 1.072 (640) | 1.047 (538) | 1.136 (77) | 26.03 | **$1.28\times10^{-5}$** | 92.3 | 26.22 | **$4.92\times10^{-5}$** | 88.6 | consistent_HET |

| | | Per-ancestry effect — OR (case N) | | | | Heterogeneity: 3 ancestries (EUR/AFR/AMR) | | | Heterogeneity: 4 ancestries (EUR/AFR/AMR/EAS) | | | |
|---|---|---|---|---|---|---|---|---|---|---|---|---|
| **Phecode** | **Description** | **EUR** | **AFR** | **AMR** | **EAS** | ***Q*** | ***p*-FDR** | ***I*² (%)** | ***Q*** | ***p*-FDR** | ***I*² (%)** | **Status** |
| NS_328 | Dementias and cerebral degeneration | 1.329 (3,212) | 1.196 (755) | 1.109 (756) | 1.184 (81) | 23.63 | **$3.41\times10^{-5}$** | 91.5 | 24.01 | **$1.15\times10^{-4}$** | 87.5 | consistent_HET |
| NS_329 | Symptoms and signs involving cognitive functions and awareness | 1.105 (19,734) | 1.037 (3,911) | 1.051 (4,161) | 1.070 (489) | 19.60 | **$2.12\times10^{-4}$** | 89.8 | 19.68 | **$7.57\times10^{-4}$** | 84.8 | consistent_HET |
| EM_239.1 | Hypercholesterolemia | 1.132 (55,789) | 1.109 (10,382) | 1.075 (11,066) | 1.155 (1,630) | 15.81 | **$1.21\times10^{-3}$** | 87.3 | 16.71 | **$2.66\times10^{-3}$** | 82.0 | consistent_HET |
| GI_542 | Chronic liver disease | 0.941 (15,294) | 0.990 (3,453) | 0.953 (6,879) | 0.855 (682) | 8.86 | **$3.42\times10^{-2}$** | 77.4 | 14.28 | **$7.33\times10^{-3}$** | 79.0 | consistent_HET |
| GI_542.2 | Fatty liver disease (FLD) | 0.930 (10,667) | 0.988 (1,947) | 0.929 (4,873) | 0.877 (528) | 8.28 | **$3.97\times10^{-2}$** | 75.9 | 9.98 | **$4.79\times10^{-2}$** | 69.9 | consistent_HET |
| EM_239.11 | Pure hypercholesterolemia | 1.136 (32,528) | 1.122 (6,075) | 1.084 (5,979) | 1.142 (857) | 8.12 | **$3.97\times10^{-2}$** | 75.4 | 8.21 | $8.08\times10^{-2}$ | 63.5 | lost_by_EAS |
| EM_239.3 | Mixed hyperlipidemia | 1.087 (31,401) | 1.086 (5,707) | 1.046 (6,529) | 1.153 (978) | 6.26 | $9.13\times10^{-2}$ | 68.1 | 9.18 | $6.20\times10^{-2}$ | 67.3 | consistent_null |
| CV_433 | Other cerebrovascular disease | 1.052 (10,138) | 1.009 (2,764) | 1.043 (1,722) | 1.203 (199) | 4.53 | $1.84\times10^{-1}$ | 55.8 | 8.19 | $8.08\times10^{-2}$ | 63.4 | consistent_null |
| EM_239.2 | Hyperglyceridemia | 1.074 (35,039) | 1.081 (6,188) | 1.041 (7,862) | 1.116 (1,160) | 5.07 | $1.52\times10^{-1}$ | 60.6 | 6.45 | $1.62\times10^{-1}$ | 53.5 | consistent_null |
| SO_374.511 | Nonexuadative (dry) age-related macular degeneration | 0.941 (3,406) | 0.937 (171) | 0.800 (234) | 0.815 (54) | 3.69 | $2.60\times10^{-1}$ | 45.7 | 4.31 | $3.78\times10^{-1}$ | 30.3 | consistent_null |
| GI_542.7 | Chronic nonalcoholic liver disease | 0.952 (6,211) | 0.993 (1,499) | 0.951 (2,910) | 0.892 (293) | 2.97 | $3.47\times10^{-1}$ | 32.7 | 4.09 | $3.86\times10^{-1}$ | 26.7 | consistent_null |
| EM_244 | Disorders of lipoprotein metabolism and other lipidemias | 1.094 (3,315) | 1.034 (388) | 1.066 (386) | 1.304 (55) | 1.60 | $5.74\times10^{-1}$ | 0.0 | 3.52 | $4.32\times10^{-1}$ | 14.8 | consistent_null |
| GE_965 | Inborn error of lipid metabolism | 1.096 (1,828) | 1.070 (205) | 0.974 (243) | 1.229 (52) | 2.69 | $3.76\times10^{-1}$ | 25.5 | 3.51 | $4.32\times10^{-1}$ | 14.6 | consistent_null |
| NS_329.8 | Altered mental status, unspecified | 1.074 (4,543) | 1.035 (1,283) | 1.059 (1,121) | 1.216 (57) | 1.86 | $5.34\times10^{-1}$ | 0.0 | 2.91 | $5.18\times10^{-1}$ | 0.0 | consistent_null |
| CV_433.2 | Occlusion and stenosis of precerebral arteries | 1.055 (6,091) | 1.043 (1,153) | 1.071 (784) | 1.212 (99) | 0.36 | $9.15\times10^{-1}$ | 0.0 | 2.08 | $6.46\times10^{-1}$ | 0.0 | consistent_null |
| CV_404 | Ischemic heart disease | 1.044 (30,905) | 1.028 (7,377) | 1.040 (5,549) | 0.997 (571) | 1.37 | $6.10\times10^{-1}$ | 0.0 | 2.05 | $6.46\times10^{-1}$ | 0.0 | consistent_null |
| CV_436 | Atherosclerosis [ASCVD] | 1.044 (32,251) | 1.033 (6,971) | 1.030 (5,447) | 1.003 (648) | 1.05 | $6.81\times10^{-1}$ | 0.0 | 1.61 | $7.21\times10^{-1}$ | 0.0 | consistent_null |
| CV_404.2 | Coronary atherosclerosis [Atherosclerotic heart disease] | 1.049 (28,166) | 1.043 (5,918) | 1.043 (4,644) | 1.020 (509) | 0.22 | $9.20\times10^{-1}$ | 0.0 | 0.45 | $9.56\times10^{-1}$ | 0.0 | consistent_null |

| | | Per-ancestry effect — OR (case N) | | | | Heterogeneity: 3 ancestries (EUR/AFR/AMR) | | | Heterogeneity: 4 ancestries (EUR/AFR/AMR/EAS) | | | |
|---|---|---|---|---|---|---|---|---|---|---|---|---|
| **Phecode** | **Description** | **EUR** | **AFR** | **AMR** | **EAS** | ***Q*** | ***p*-FDR** | **$I^2$ (%)** | ***Q*** | ***p*-FDR** | **$I^2$ (%)** | **Status** |
| CV_403 | Angina pectoris | 1.049 (9,769) | 1.040 (2,339) | 1.044 (1,866) | 1.009 (151) | 0.17 | $9.20\times10^{-1}$ | 0.0 | 0.32 | $9.56\times10^{-1}$ | 0.0 | consistent_null |

Cochran's Q test for ancestry heterogeneity across the 9 phecodes with FDR-significant heterogeneity in the primary 4-ancestry analysis (EUR, AFR, AMR, EAS). To assess whether the smaller East Asian sample (n = 9,549) was driving the observed heterogeneity, we repeated the test excluding EAS (3-ancestry analysis: EUR, AFR, AMR). Cochran's Q p-values are shown for each analysis alongside Benjamini–Hochberg FDR-adjusted p-values. Phecodes retaining FDR-significant heterogeneity (FDR-p < 0.05) in both analyses are marked as concordant; these are considered robust ancestry-heterogeneous signals. All estimates are from ancestry-stratified ordinal APOE logistic regressions adjusted for age, sex at birth, EHR length, and 16 genetic principal components (Methods).

**Table S11.** Plasma proteins reaching FDR significance for APOE ε4 dosage in the whole cohort, adjusted for ε2 dosage.

| Protein | N | β (SE) | P | P (FDR) |
|---|---|---|---|---|
| **MENT** | 9,115 | +0.607 (0.020) | 1.37e-197 | 7.41e-194 |
| **SNAP25** | 9,123 | +0.589 (0.020) | 1.35e-186 | 3.64e-183 |
| **PALM** | 9,088 | +0.218 (0.021) | 4.75e-26 | 8.57e-23 |
| **CSNK2A1** | 9,126 | +0.204 (0.021) | 1.23e-21 | 1.67e-18 |
| **PLA2G7** | 9,101 | +0.188 (0.021) | 1.23e-18 | 1.33e-15 |
| **BRK1** | 9,112 | +0.154 (0.019) | 1.75e-16 | 1.58e-13 |
| **RP2** | 9,115 | +0.141 (0.018) | 2.88e-15 | 2.23e-12 |
| **BPIFB2** | 9,126 | -0.135 (0.021) | 7.09e-11 | 4.80e-08 |
| **HMOX2** | 9,094 | +0.106 (0.018) | 8.17e-09 | 4.91e-06 |
| **ANKS1A** | 9,126 | +0.091 (0.016) | 3.47e-08 | 1.88e-05 |
| **SMPDL3A** | 9,116 | -0.119 (0.022) | 4.61e-08 | 2.27e-05 |
| **AGR3** | 9,131 | +0.107 (0.021) | 6.35e-07 | 2.87e-04 |
| **IDUA** | 9,119 | -0.097 (0.021) | 6.79e-06 | 2.83e-03 |
| **APOF** | 9,098 | +0.083 (0.019) | 9.93e-06 | 3.84e-03 |
| **APOA2** | 9,103 | +0.090 (0.021) | 2.57e-05 | 9.29e-03 |
| **CRISP2** | 9,115 | +0.081 (0.020) | 3.39e-05 | 1.15e-02 |
| **CES1** | 9,113 | -0.084 (0.021) | 6.15e-05 | 1.96e-02 |

Fourteen proteins reached Benjamini–Hochberg FDR significance (P (FDR) < 0.05) in a joint linear model with additive ε4 allele count as exposure, adjusted for ε2 dosage in addition to the standard covariates. Rows are sorted by ascending raw P value. Sample size (N) reflects the number of participants with non-missing measurements for each protein. β, regression coefficient per additional ε4 allele on the standardised protein level; SE, standard error.

**Table S12.** Plasma proteins reaching FDR significance for APOE ε2 dosage in the whole cohort, adjusted for ε4 dosage.

| Protein | N | β (SE) | P | P (FDR) |
|---|---|---|---|---|
| **MENT** | 9,115 | +0.392 (0.026) | 1.60e-51 | 8.66e-48 |
| **LDLR** | 9,112 | +0.396 (0.028) | 4.04e-45 | 1.09e-41 |
| **PLA2G7** | 9,101 | -0.373 (0.028) | 1.38e-40 | 2.50e-37 |
| **BRK1** | 9,112 | -0.325 (0.024) | 3.76e-40 | 5.09e-37 |
| **FGFBP1** | 9,086 | -0.202 (0.029) | 2.74e-12 | 2.97e-09 |
| **HMOX1** | 9,110 | +0.182 (0.027) | 3.07e-11 | 2.77e-08 |
| **HEXIM1** | 9,128 | -0.146 (0.023) | 3.86e-10 | 2.99e-07 |
| **PDGFC** | 9,122 | +0.157 (0.028) | 2.79e-08 | 1.89e-05 |
| **UBL4A** | 9,116 | +0.126 (0.023) | 4.79e-08 | 2.88e-05 |
| **CCN1** | 9,126 | +0.143 (0.026) | 6.71e-08 | 3.48e-05 |
| **AGRP** | 9,127 | +0.153 (0.028) | 7.08e-08 | 3.48e-05 |
| **ANGPTL3** | 9,097 | +0.144 (0.027) | 1.09e-07 | 4.91e-05 |
| **NPY** | 9,128 | +0.148 (0.028) | 1.71e-07 | 7.14e-05 |
| **CCK** | 9,130 | +0.146 (0.028) | 2.65e-07 | 1.02e-04 |
| **SORT1** | 9,115 | +0.127 (0.025) | 2.88e-07 | 1.04e-04 |
| **DEFB4A_DEFB4B** | 9,132 | +0.139 (0.027) | 3.97e-07 | 1.34e-04 |
| **PBXIP1** | 9,113 | +0.136 (0.027) | 5.25e-07 | 1.67e-04 |
| **LPA** | 9,116 | -0.132 (0.028) | 1.78e-06 | 5.34e-04 |
| **PLA2G10** | 9,122 | +0.131 (0.028) | 2.35e-06 | 6.69e-04 |
| **CELA2A** | 9,105 | +0.130 (0.028) | 4.80e-06 | 1.30e-03 |
| **PTH** | 9,120 | +0.126 (0.028) | 6.47e-06 | 1.67e-03 |
| **EXTL1** | 9,103 | +0.130 (0.029) | 8.27e-06 | 2.04e-03 |
| **TNFSF13** | 9,100 | +0.108 (0.025) | 1.18e-05 | 2.77e-03 |
| **CCL18** | 9,102 | +0.110 (0.026) | 2.46e-05 | 5.54e-03 |
| **APOD** | 9,088 | +0.117 (0.028) | 3.08e-05 | 6.67e-03 |
| **IL6ST** | 9,113 | +0.110 (0.027) | 4.69e-05 | 9.77e-03 |

| Protein | N | β (SE) | P | P (FDR) |
|---|---|---|---|---|
| **KRT5** | 9,126 | +0.112 (0.028) | 5.00e-05 | 1.00e-02 |
| **ADGRF5** | 9,116 | +0.108 (0.027) | 5.61e-05 | 1.08e-02 |
| **LENG1** | 9,124 | +0.083 (0.021) | 6.59e-05 | 1.22e-02 |
| **LENG8** | 9,127 | -0.106 (0.027) | 6.76e-05 | 1.22e-02 |
| **SERPINI2** | 9,127 | +0.114 (0.029) | 7.07e-05 | 1.22e-02 |
| **APOA1** | 9,095 | +0.110 (0.028) | 7.27e-05 | 1.22e-02 |
| **CCL27** | 9,117 | +0.095 (0.024) | 7.41e-05 | 1.22e-02 |
| **SEMA3G** | 9,104 | +0.112 (0.029) | 8.62e-05 | 1.37e-02 |
| **GPNMB** | 9,117 | +0.105 (0.027) | 9.81e-05 | 1.52e-02 |
| **CA14** | 9,109 | +0.108 (0.028) | 1.01e-04 | 1.53e-02 |
| **BGLAP** | 9,118 | -0.108 (0.028) | 1.26e-04 | 1.84e-02 |
| **CPA2** | 9,116 | +0.108 (0.028) | 1.34e-04 | 1.88e-02 |
| **GRP** | 9,123 | +0.107 (0.028) | 1.35e-04 | 1.88e-02 |
| **VMO1** | 9,114 | +0.102 (0.027) | 1.41e-04 | 1.91e-02 |
| **LHPP** | 9,111 | +0.100 (0.026) | 1.45e-04 | 1.91e-02 |
| **EGFL6** | 9,122 | +0.101 (0.027) | 1.53e-04 | 1.97e-02 |
| **EMC7** | 9,060 | +0.109 (0.029) | 1.92e-04 | 2.42e-02 |
| **BMP10** | 9,113 | +0.104 (0.028) | 2.05e-04 | 2.52e-02 |
| **FAM3C** | 9,115 | +0.090 (0.025) | 2.43e-04 | 2.89e-02 |
| **GPR37** | 9,123 | +0.098 (0.027) | 2.47e-04 | 2.89e-02 |
| **THBD** | 9,112 | +0.093 (0.025) | 2.51e-04 | 2.89e-02 |
| **TINAGL1** | 9,095 | +0.101 (0.028) | 2.69e-04 | 3.04e-02 |
| **PRR32** | 9,132 | -0.105 (0.029) | 2.75e-04 | 3.04e-02 |
| **MANSC1** | 9,113 | +0.102 (0.028) | 2.89e-04 | 3.13e-02 |
| **CD300A** | 9,113 | +0.094 (0.026) | 2.97e-04 | 3.13e-02 |
| **S100P** | 9,121 | +0.099 (0.027) | 3.05e-04 | 3.13e-02 |
| **PPP2R5A** | 9,122 | -0.085 (0.024) | 3.11e-04 | 3.13e-02 |
| **MYH10** | 9,124 | +0.097 (0.027) | 3.12e-04 | 3.13e-02 |
| **RGMA** | 9,116 | +0.102 (0.028) | 3.21e-04 | 3.14e-02 |
| **VCAN** | 9,114 | +0.099 (0.028) | 3.24e-04 | 3.14e-02 |

| Protein | N | β (SE) | P | P (FDR) |
|---|---|---|---|---|
| **FGFBP2** | 9,112 | +0.101 (0.028) | 3.31e-04 | 3.15e-02 |
| **GRN** | 9,100 | +0.100 (0.028) | 3.38e-04 | 3.16e-02 |
| **PDCD5** | 9,109 | +0.080 (0.022) | 3.66e-04 | 3.25e-02 |
| **ST3GAL1** | 9,114 | +0.100 (0.028) | 3.67e-04 | 3.25e-02 |
| **LMOD1** | 9,080 | +0.078 (0.022) | 3.68e-04 | 3.25e-02 |
| **CD34** | 9,115 | +0.103 (0.029) | 3.72e-04 | 3.25e-02 |
| **SOD1** | 9,084 | +0.087 (0.025) | 4.22e-04 | 3.54e-02 |
| **SWAP70** | 9,105 | +0.089 (0.025) | 4.23e-04 | 3.54e-02 |
| **TNFSF10** | 9,120 | +0.100 (0.028) | 4.25e-04 | 3.54e-02 |
| **CEP290** | 9,117 | -0.097 (0.028) | 5.01e-04 | 4.07e-02 |
| **IL18** | 9,099 | +0.093 (0.027) | 5.04e-04 | 4.07e-02 |
| **GP5** | 9,112 | +0.094 (0.027) | 5.25e-04 | 4.12e-02 |
| **LGMN** | 9,117 | +0.095 (0.027) | 5.31e-04 | 4.12e-02 |
| **PLA2G1B** | 9,098 | +0.100 (0.029) | 5.32e-04 | 4.12e-02 |
| **DNAJB1** | 9,125 | +0.079 (0.023) | 5.40e-04 | 4.12e-02 |
| **KLF4** | 9,121 | +0.095 (0.027) | 5.77e-04 | 4.34e-02 |
| **EIF4EBP1** | 9,108 | +0.085 (0.025) | 6.08e-04 | 4.51e-02 |
| **ENTPD5** | 9,121 | +0.098 (0.029) | 6.26e-04 | 4.55e-02 |
| **CELSR2** | 9,116 | +0.098 (0.029) | 6.30e-04 | 4.55e-02 |
| **NID1** | 9,094 | +0.090 (0.027) | 6.60e-04 | 4.70e-02 |
| **NTRK3** | 9,117 | +0.090 (0.027) | 7.02e-04 | 4.94e-02 |

List of the 77 proteins that reached Benjamini–Hochberg FDR significance (P (FDR) < 0.05) in a joint linear model with additive ε2 allele count as exposure, adjusted for ε4 dosage in addition to the standard covariates. Rows are sorted by ascending raw P value. Sample size (N) reflects the number of participants with non-missing measurements for each protein. β, regression coefficient per additional ε2 allele on the standardised protein level; SE, standard error.

**Table S13.** Ancestry-stratified plasma proteins reaching FDR significance for APOE ε2 allele dosage.

| Ancestry | Protein | N | β (SE) | P | P (FDR) |
|---|---|---|---|---|---|
| **AFR** | **BRK1** | 1,437 | -0.406 (0.055) | 4.06e-13 | 2.20e-09 |
| | **MENT** | 1,439 | +0.379 (0.061) | 7.05e-10 | 1.91e-06 |
| | **PLA2G7** | 1,436 | -0.374 (0.061) | 1.31e-09 | 2.37e-06 |

| Ancestry | Protein | N | β (SE) | P | P (FDR) |
|---|---|---|---|---|---|
| | **LDLR** | 1,438 | +0.360 (0.062) | 8.51e-09 | 1.15e-05 |
| | **PDGFC** | 1,437 | +0.258 (0.059) | 1.11e-05 | 1.05e-02 |
| | **CDC42EP1** | 1,438 | -0.262 (0.060) | 1.17e-05 | 1.05e-02 |
| **AMR** | **PLA2G7** | 2,030 | -0.431 (0.068) | 2.78e-10 | 9.79e-07 |
| | **MENT** | 2,029 | +0.428 (0.068) | 3.61e-10 | 9.79e-07 |
| | **LDLR** | 2,030 | +0.368 (0.071) | 2.57e-07 | 4.65e-04 |
| | **ITGB5** | 2,030 | +0.275 (0.064) | 1.83e-05 | 2.47e-02 |
| **EAS** | **LDLR** | 1,369 | +0.447 (0.067) | 4.56e-11 | 2.47e-07 |
| | **PLA2G7** | 1,362 | -0.457 (0.073) | 5.83e-10 | 1.58e-06 |
| | **FGFBP1** | 1,361 | -0.346 (0.064) | 7.72e-08 | 1.39e-04 |
| | **LPA** | 1,372 | -0.283 (0.061) | 4.07e-06 | 5.51e-03 |
| **EUR** | **MENT** | 2,644 | +0.510 (0.045) | 6.28e-29 | 3.40e-25 |
| | **BRK1** | 2,639 | -0.389 (0.041) | 1.17e-20 | 3.17e-17 |
| | **LDLR** | 2,643 | +0.397 (0.048) | 2.08e-16 | 3.75e-13 |
| | **PLA2G7** | 2,642 | -0.323 (0.048) | 1.45e-11 | 1.96e-08 |
| | **HMOX1** | 2,642 | +0.223 (0.046) | 1.58e-06 | 1.71e-03 |
| | **APOA1** | 2,638 | +0.204 (0.049) | 3.25e-05 | 2.93e-02 |
| | **TCTN3** | 2,643 | +0.212 (0.052) | 4.89e-05 | 3.78e-02 |
| | **HEXIM1** | 2,648 | -0.168 (0.042) | 6.40e-05 | 4.33e-02 |
| **SAS** | **LDLR** | 1,185 | +0.508 (0.088) | 1.11e-08 | 6.03e-05 |
| | **BRK1** | 1,186 | -0.399 (0.077) | 2.77e-07 | 7.50e-04 |

Shown are the 24 ancestry × protein tests (across 13 unique proteins) reaching Benjamini–Hochberg FDR significance (P (FDR) < 0.05) for APOE ε2 allele dosage. FDR was computed within each ancestry over 5,415 tested proteins. Effect sizes come from a joint linear model fit separately within each ancestry, in which the ε2 coefficient is adjusted for ε4 dosage in addition to the standard covariates. Rows are grouped by ancestry and, within each ancestry, sorted by ascending raw P value. AFR, African/African American; AMR, Admixed American; EAS, East Asian; EUR, European; SAS, South Asian. β, regression coefficient per additional ε2 allele on the standardised protein level; SE, standard error.

**Table S14.** Ancestry-stratified plasma proteins reaching FDR significance for APOE ε4 allele dosage.

| Ancestry | Protein | N | β (SE) | P | P (FDR) |
|---|---|---|---|---|---|
| **AFR** | **SNAP25** | 1,438 | +0.519 (0.042) | 1.34e-33 | 7.24e-30 |
| | **MENT** | 1,439 | +0.542 (0.045) | 1.12e-31 | 3.04e-28 |

| Ancestry | Protein | N | β (SE) | P | P (FDR) |
|---|---|---|---|---|---|
| | **CSNK2A1** | 1,438 | +0.272 (0.045) | 2.85e-09 | 5.14e-06 |
| | **PLA2G7** | 1,436 | +0.229 (0.045) | 4.69e-07 | 6.34e-04 |
| | **IL32** | 1,435 | +0.194 (0.046) | 3.08e-05 | 3.33e-02 |
| **AMR** | **MENT** | 2,029 | +0.646 (0.041) | 3.36e-52 | 1.82e-48 |
| | **SNAP25** | 2,033 | +0.537 (0.041) | 4.74e-38 | 1.28e-34 |
| | **CSNK2A1** | 2,033 | +0.184 (0.043) | 2.30e-05 | 4.15e-02 |
| **EAS** | **SNAP25** | 1,373 | +0.667 (0.056) | 8.72e-31 | 4.72e-27 |
| | **MENT** | 1,369 | +0.529 (0.053) | 1.67e-22 | 4.53e-19 |
| | **PALM** | 1,368 | +0.338 (0.059) | 1.09e-08 | 1.97e-05 |
| **EUR** | **MENT** | 2,644 | +0.651 (0.037) | 1.88e-65 | 1.02e-61 |
| | **SNAP25** | 2,643 | +0.595 (0.037) | 3.82e-55 | 1.04e-51 |
| | **PALM** | 2,638 | +0.242 (0.037) | 1.10e-10 | 1.98e-07 |
| | **BRK1** | 2,639 | +0.211 (0.034) | 5.36e-10 | 7.25e-07 |
| | **RP2** | 2,643 | +0.204 (0.035) | 5.05e-09 | 5.27e-06 |
| | **CSNK2A1** | 2,644 | +0.233 (0.040) | 5.84e-09 | 5.27e-06 |
| | **ANKS1A** | 2,645 | +0.154 (0.032) | 1.10e-06 | 8.51e-04 |
| | **SMPD1** | 2,643 | -0.191 (0.039) | 1.34e-06 | 9.10e-04 |
| | **BMERB1** | 2,630 | +0.185 (0.040) | 3.07e-06 | 1.85e-03 |
| | **SMPDL3A** | 2,644 | -0.184 (0.042) | 1.33e-05 | 7.22e-03 |
| | **APOF** | 2,641 | +0.133 (0.034) | 7.32e-05 | 3.35e-02 |
| | **CLIP2** | 2,647 | +0.144 (0.036) | 7.97e-05 | 3.35e-02 |
| | **CREG1** | 2,642 | -0.156 (0.040) | 8.04e-05 | 3.35e-02 |
| **SAS** | **SNAP25** | 1,187 | +0.706 (0.060) | 2.08e-30 | 1.13e-26 |
| | **MENT** | 1,185 | +0.611 (0.056) | 1.19e-26 | 3.22e-23 |
| | **PALM** | 1,184 | +0.267 (0.056) | 1.89e-06 | 3.41e-03 |

Shown are the 27 ancestry × protein tests (across 15 unique proteins) reaching Benjamini–Hochberg FDR significance (P (FDR) < 0.05) for APOE ε4 allele dosage. FDR was computed within each ancestry over 5,415 tested proteins. Effect sizes come from a joint linear model fit separately within each ancestry, in which the ε4 coefficient is adjusted for ε2 dosage in addition to the standard covariates. Rows are grouped by ancestry and, within each ancestry, sorted by ascending raw P value. AFR, African/African American; AMR, Admixed American; EAS, East Asian; EUR, European; SAS, South Asian. β, regression coefficient per additional ε4 allele on the standardised protein level; SE, standard error.

**Table S15.** Plasma SNAP25 levels in participants with dementia-related phecodes versus controls.

| Phecode | Phenotype | Cases | Controls | β (95% CI) | P | P (FDR) |
|---|---|---|---|---|---|---|
| NS_329.1 | Memory loss | 249 | 6,709 | +0.200 (+0.079, +0.320) | 1.17e-03 | 4.69e-03 |
| NS_328.11 | Alzheimer's disease | 28 | 7,064 | +0.476 (+0.128, +0.824) | 7.40e-03 | 0.015 |
| NS_329.5 | Mild cognitive impairment | 103 | 6,957 | +0.151 (-0.032, +0.335) | 0.106 | 0.142 |
| NS_328.1 | Dementias | 105 | 6,948 | +0.119 (-0.063, +0.302) | 0.200 | 0.200 |

Case-control comparison of plasma SNAP25 inverse rank normalized NPX value within the proteomic subcohort (n = 7,104), fitted as an adjusted linear regression (SNAP25 ~ case + age + sex at birth + EHR length + mean NPX + plate ID + 16 genetic principal components). Cases were defined as ≥2 occurrences of the phecode; controls as zero occurrences. β represents the adjusted mean difference in SNAP25 NPX (case − control) with 95% confidence interval. Phecodes are ordered from most to least clinically specific (Alzheimer's disease → Memory loss). Raw P values and Benjamini–Hochberg FDR-adjusted P values across the four pre-specified phecodes are reported.

### Supplementary Figures

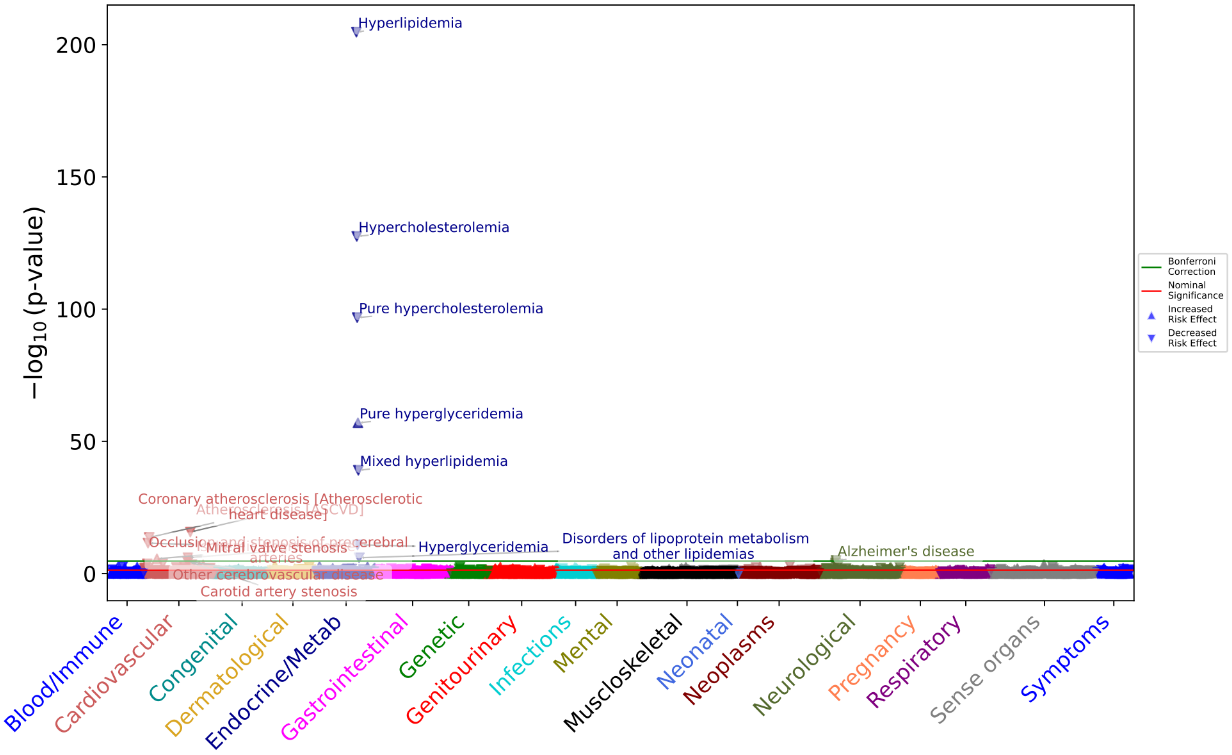


**Figure S1.** PheWAS analysis of APOE ε2 allele dosage in the whole cohort (n = 367,757), joint model. Logistic regressions for each phecode were fit with an additive ε2 allele count (0/1/2) as the exposure, adjusted for ε4 allele dosage in addition to age, sex, 16 genetic PCs and EHR length. A total of 2,627 phecodes were evaluated. The green line represents the significance threshold after Bonferroni correction ($p < 1.90\times10^{-5}$). The red line represents the nominal significance threshold of 0.05. Triangles pointing up indicate that additional ε2 copies increase the odds of the phecode (OR > 1); inverted triangles indicate that additional ε2 copies decrease the odds (OR < 1). X-axis shows phecodes aggregated into 18 categories, as implemented by the PheTK package. Y-axis displays the -$\log_{10}$(p-value) for each phecode.

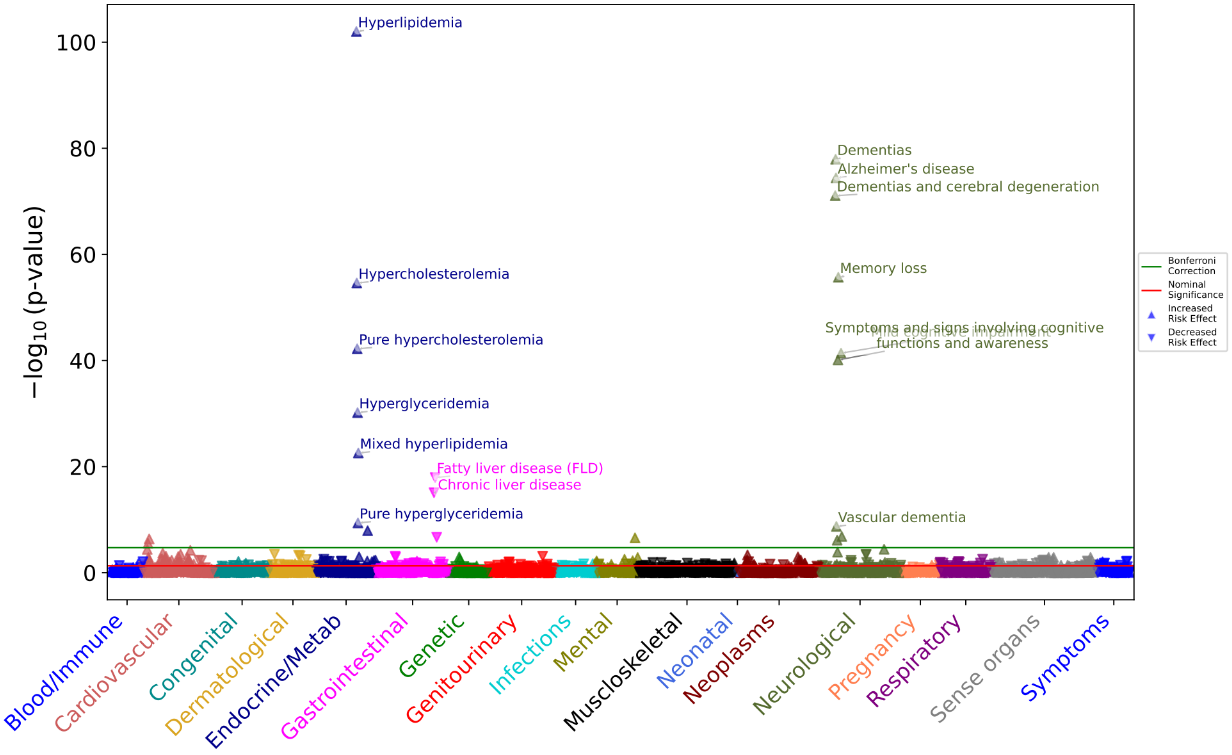


**Figure S2.** PheWAS analysis of APOE ε4 allele dosage in the whole cohort (n = 367,757), joint model. Logistic regressions for each phecode were fit with an additive ε4 allele count (0/1/2) as the exposure, adjusted for ε2 allele dosage in addition to age, sex, 16 genetic PCs and EHR length. A total of 2,627 phecodes were evaluated. The green line represents the significance threshold after Bonferroni correction ($p < 1.90\times10^{-5}$). The red line represents the nominal significance threshold of 0.05. Triangles pointing up indicate that additional ε4 copies increase the odds of the phecode (OR > 1); inverted triangles indicate that additional ε4 copies decrease the odds (OR < 1). The x-axis shows phecodes aggregated into 18 categories, as implemented by the PheTK package. Y-axis displays the $-\log_{10}$(p-value) for each phecode.

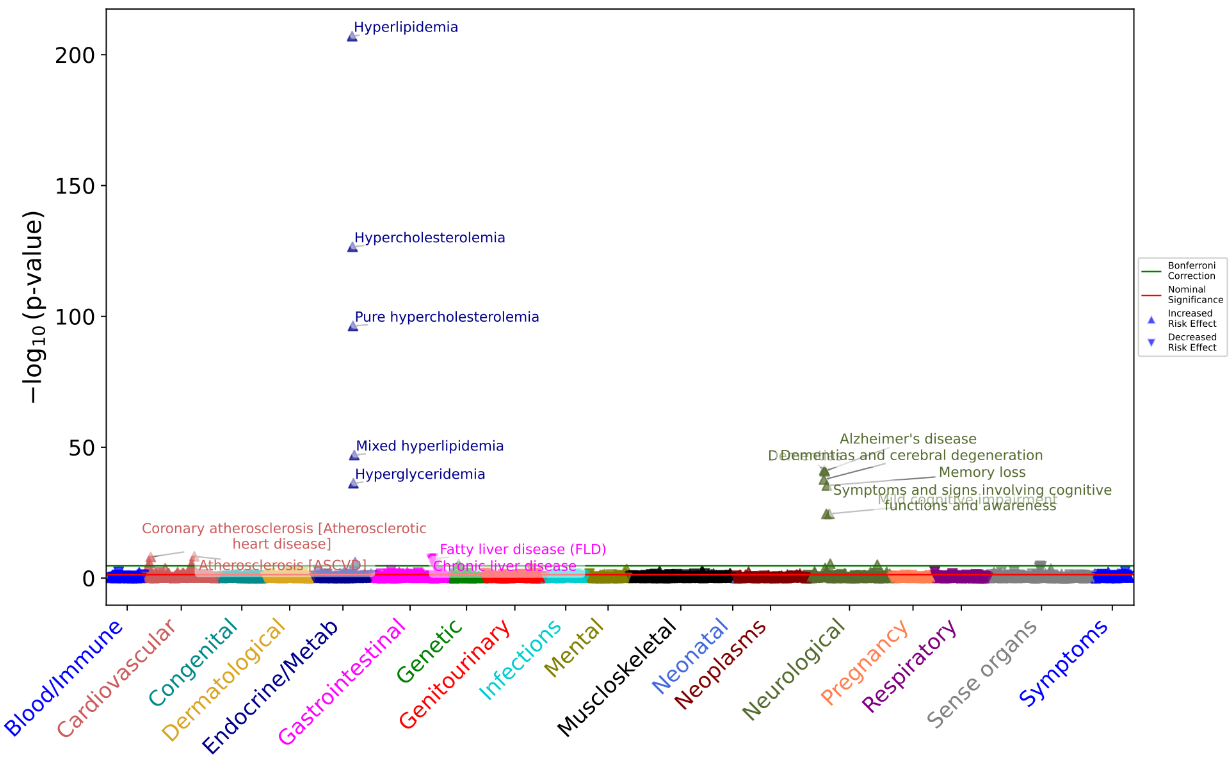


**Figure S3.** PheWAS analysis of APOE genotypes in female participants (sex assigned at birth; n=224,916). Genotypes were arranged hierarchically from ε2 to ε4 (ε2/ε2 < ε2/ε3 < ε3/ε3 < ε2/ε4 < ε3/ε4 < ε4/ε4). Logistic regressions for each phecode were adjusted by age, 16 genetic PCs and EHR length (sex omitted from the covariate set given the stratification). A total of 2,384 phecodes were evaluated. The green line represents the stratum-specific Bonferroni-corrected significance threshold ($p < 2.10\times10^{-5}$). The red line represents the nominal significance threshold of 0.05. Triangles pointing up indicate a OR > 1; inverted triangles indicate a OR < 1. The x-axis shows phecodes aggregated into 18 categories, as implemented by the PheTK package. Y-axis displays the -$\log_{10}$(p-value) for each phecode.

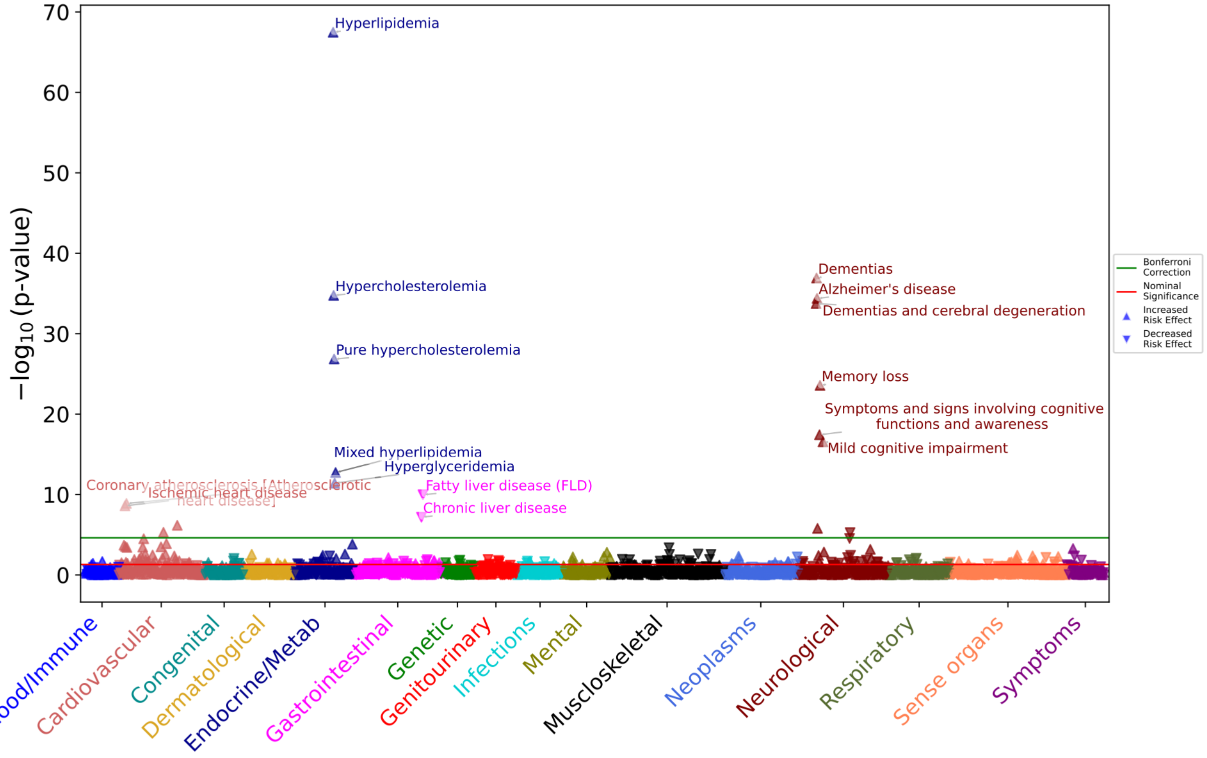


**Figure S4.** PheWAS analysis of APOE genotypes in male participants (sex assigned at birth; n = 142,841). Genotypes were arranged hierarchically from ε2 to ε4 (ε2/ε2 < ε2/ε3 < ε3/ε3 < ε2/ε4 < ε3/ε4 < ε4/ε4). Logistic regressions for each phecode were adjusted by age, 16 genetic PCs and EHR length (sex omitted from the covariate set given the stratification). A total of 2,066 phecodes were evaluated. The green line represents the stratum-specific Bonferroni-corrected significance threshold ($p < 2.42\times10^{-5}$). The red line represents the nominal significance threshold of 0.05. Triangles pointing up indicate a OR > 1; inverted triangles indicate a OR < 1. The x-axis shows phecodes aggregated into 18 categories, as implemented by the PheTK package. The y-axis displays the $-\log_{10}$(p-value) for each phecode.

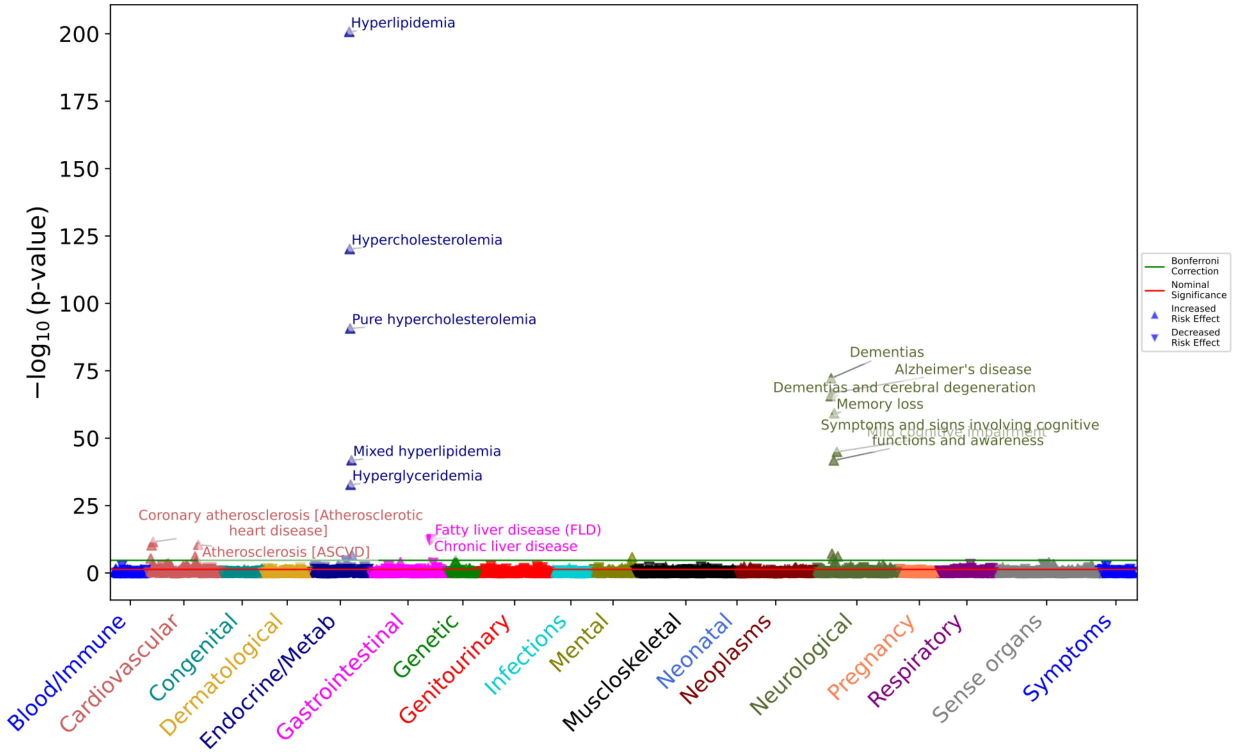


**Figure S5.** PheWAS analysis of APOE genotypes in participants of European ancestry (EUR; n=221,901). Genotypes were arranged hierarchically from ε2 to ε4 (ε2/ε2 < ε2/ε3 < ε3/ε3 < ε2/ε4 < ε3/ε4 < ε4/ε4). Logistic regressions for each phecode were adjusted by age, sex, 16 genetic PCs and EHR length. A total of 2,272 phecodes were evaluated. The green line represents the ancestry-specific Bonferroni-corrected significance threshold ($p < 2.20\times10^{-5}$). The red line represents the nominal significance threshold of 0.05. Triangles pointing up indicate a OR > 1; inverted triangles indicate a OR < 1. The x-axis shows phecodes aggregated into 18 categories, as implemented by the PheTK package. Y-axis displays the $-\log_{10}$(p-value) for each phecode.

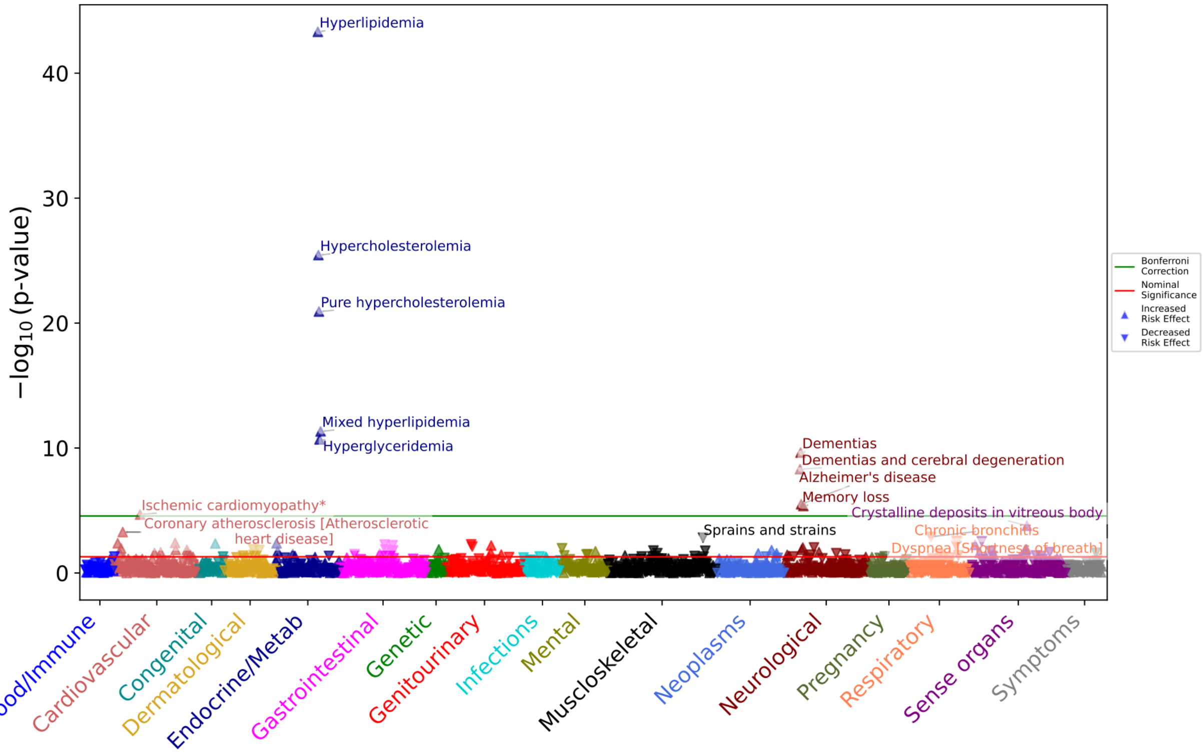


**Figure S6.** PheWAS analysis of APOE genotypes in participants of African ancestry (AFR; n=64,261). Genotypes were arranged hierarchically from ε2 to ε4 (ε2/ε2 < ε2/ε3 < ε3/ε3 < ε2/ε4 < ε3/ε4 < ε4/ε4). Logistic regressions for each phecode were adjusted by age, sex, 16 genetic PCs and EHR length. A total of 1,824 phecodes were evaluated. The green line represents the ancestry-specific Bonferroni-corrected significance threshold ($p < 2.74\times10^{-5}$). The red line represents the nominal significance threshold of 0.05. Triangles pointing up indicate a OR > 1; inverted triangles indicate a OR < 1. The x-axis shows phecodes aggregated into 18 categories, as implemented by the PheTK package. Y-axis displays the $-\log_{10}$(p-value) for each phecode.

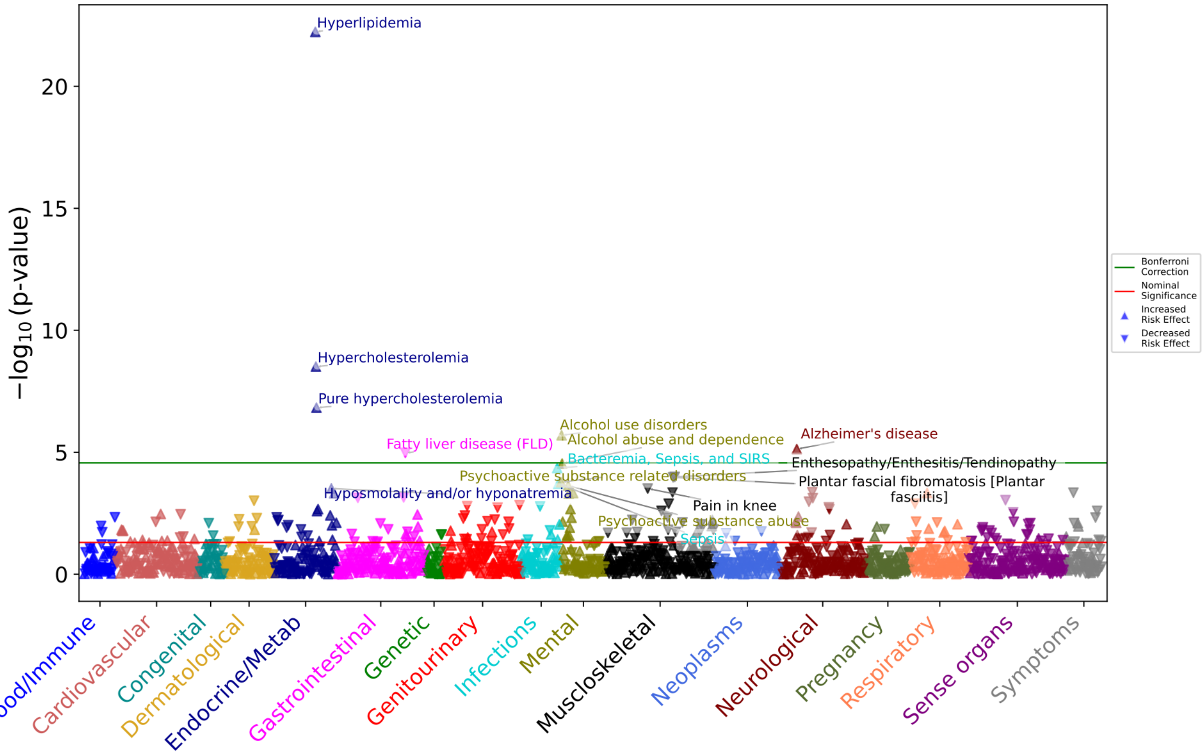


**Figure S7.** PheWAS analysis of APOE genotypes in participants of Admixed American ancestry (AMR; n=66,856). Genotypes were arranged hierarchically from ε2 to ε4 (ε2/ε2 < ε2/ε3 < ε3/ε3 < ε2/ε4 < ε3/ε4 < ε4/ε4). Logistic regressions for each phecode were adjusted by age, sex, 16 genetic PCs and EHR length. A total of 1,843 phecodes were evaluated. The green line represents the ancestry-specific Bonferroni-corrected significance threshold ($p < 2.71\times10^{-5}$). The red line represents the nominal significance threshold of 0.05. Triangles pointing up indicate a OR > 1; inverted triangles indicate a OR < 1. The x-axis shows phecodes aggregated into 18 categories, as implemented by the PheTK package. Y-axis displays the $-\log_{10}$(p-value) for each phecode.

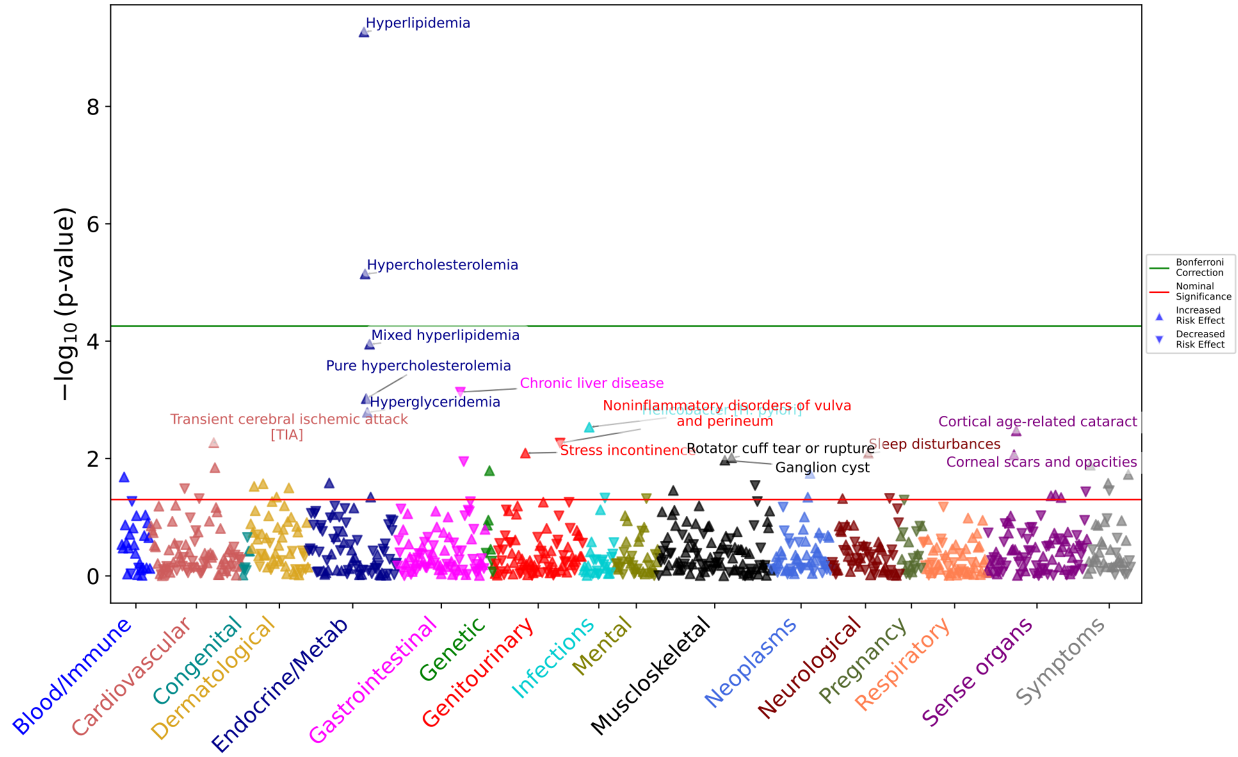


**Figure S8.** PheWAS analysis of APOE genotypes in participants of East Asian ancestry (EAS; n = 9,549). Genotypes were arranged hierarchically from ε2 to ε4 (ε2/ε2 < ε2/ε3 < ε3/ε3 < ε2/ε4 < ε3/ε4 < ε4/ε4). Logistic regressions for each phecode were adjusted by age, sex, 16 genetic PCs and EHR length. A total of 901 phecodes were evaluated. The green line represents the ancestry-specific Bonferroni-corrected significance threshold ($p < 5.55\times10^{-5}$). The red line represents the nominal significance threshold of 0.05. Triangles pointing up indicate a OR > 1; inverted triangles indicate a OR < 1. The x-axis shows phecodes aggregated into 18 categories, as implemented by the PheTK package. Y-axis displays the -$\log_{10}$(p-value) for each phecode.

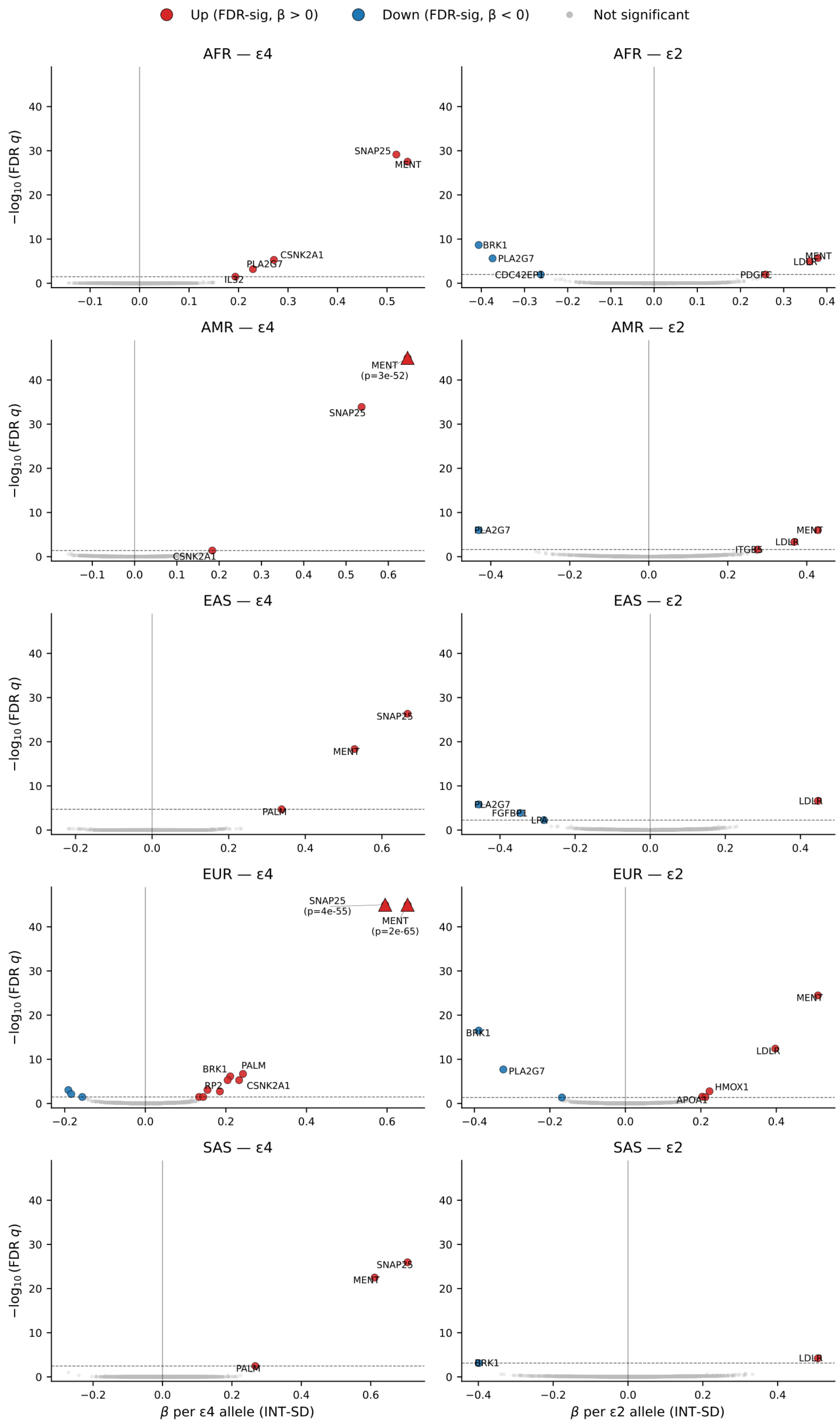

Up (FDR-sig, β > 0)
Down (FDR-sig, β < 0)
Not significant
AFR — ε4
AFR — ε2
AMR — ε4
AMR — ε2
EAS — ε4
EAS — ε2
EUR — ε4
EUR — ε2
SAS — ε4
SAS — ε2
−log10(FDR q)
SNAP25
MENT
CSNK2A1
PLA2G7
IL32
BRK1
CDC42EP1
LDLR
PDGFC
MENT (p=3e-52)
ITGB5
PALM
FGFBP1
LPA
SNAP25 (p=4e-55)
MENT (p=2e-65)
RP2
HMOX1
APOA1
β per ε4 allele (INT-SD)
β per ε2 allele (INT-SD)

**Figure S9:** Ancestry-stratified plasma pQTL volcano plots for APOE ε4 (left column) and ε2 (right column) allele dosage across five ancestral groups (rows): African/African American (AFR), Admixed American (AMR), East Asian (EAS), European (EUR), and South Asian (SAS). For each stratum, a joint linear model was fit within the ancestry, in which the ε2 coefficient is adjusted for ε4 allele dosage and the ε4 coefficient is adjusted for ε2 allele dosage, in addition to age, sex, 16 genetic PCs and EHR length. All 5,415 available Olink proteins were tested per stratum. Each dot represents one protein: red dots indicate proteins reaching Benjamini–Hochberg FDR significance (P (FDR) < 0.05) with a positive effect (β > 0), blue dots indicate FDR-significant proteins with a negative effect (β < 0), and grey dots indicate non-significant proteins. FDR was computed independently within each ancestry × allele stratum. The dashed horizontal line marks the stratum-specific FDR = 0.05 threshold; the vertical black line marks β = 0. Proteins whose FDR-adjusted $-\log_{10}$ value exceeded the y-axis cap (45) are displayed as triangles at the top of the panel with the raw P value annotated. The top six FDR-significant proteins per panel are labeled with gene symbols. X-axis, regression coefficient (β) per additional allele on the standardised protein level; Y-axis, $-\log_{10}$ of the Benjamini–Hochberg FDR-adjusted P value.
